# Machine learning in the identification of prognostic DNA methylation biomarkers among patients with cancer: a systematic review of epigenome-wide studies

**DOI:** 10.1101/2022.09.02.22279533

**Authors:** Tanwei Yuan, Dominic Edelmann, Ziwen Fan, Elizabeth Alwers, Jakob Nikolas Kather, Hermann Brenner, Michael Hoffmeister

## Abstract

**Summary:** *Background:* DNA methylation biomarkers have great potential in improving prognostic classification systems for patients with cancer. Machine learning (ML)-based analytic techniques might help overcome the challenges of analyzing high-dimensional data in relatively small sample sizes. This systematic review summarizes the current use of ML-based methods in epigenome-wide studies for the identification of DNA methylation signatures associated with cancer prognosis.

*Methods:* We searched three electronic databases including PubMed, EMBASE, and Web of Science for articles published until 8 June 2022. ML-based methods and workflows used to identify DNA methylation signatures associated with cancer prognosis were extracted and summarized. Two authors independently assessed the methodological quality of included studies by a seven-item checklist adapted from relevant guidelines.

*Results:* Seventy-six studies were included in this review. Three major types of ML-based workflows were identified: 1) unsupervised clustering, 2) supervised feature selection, and 3) deep learning-based feature transformation. For the three workflows, the most frequently used ML techniques were consensus clustering, least absolute shrinkage and selection operator (LASSO), and autoencoder, respectively. The systematic review revealed that the performance of these approaches has not been adequately evaluated yet and that methodological and reporting flaws were common in the identified studies using ML techniques.

*Conclusions:* There is great heterogeneity in ML-based methodological strategies used by epigenome-wide studies to identify DNA methylation markers associated with cancer prognosis. Benchmarking studies are needed to compare the relative performance of various approaches for specific cancer types. Adherence to relevant methodological and reporting guidelines is urgently needed.

## Background

Cancer remains a leading cause of death and is projected to become even more frequent due to population aging and growth in nearly every country of the world (1). Accurate prediction of prognosis for patients with cancer is pivotal for individualized treatment and reduction in mortality. So far, the tumor-lymph node-metastasis (TNM) staging system has been the most commonly used system for treatment decisions and to predict the prognosis of cancer (2-4). However, the role of molecular features is becoming increasingly important for a more precise treatment and prediction of cancer prognosis (3, 4).

DNA methylation involves the addition of a methyl group to the C5 position of the cytosine to form 5-methylcytosine (5). It is one of the most common epigenetic changes that regulate gene expression and plays a key role in carcinogenesis, cancer development, and clinical prognosis (5). DNA methylation signatures hold great promise for improving the prognostic accuracy of different cancers (6-9). Recent advances in DNA methylation microarray platforms enable studies to analyze methylation across the whole genome in a high-throughput manner (10, 11). However, the sample size of these epigenome-wide studies is often less than 1000 patients, much smaller relative to the number of CpG sites investigated (27,578-850,000). Such analyses are prone to the issues of overfitting and multicollinearity when using traditional statistical methods, most of which are intended for the low-dimensional setting (12).

Machine learning (ML) could be a powerful tool to properly handle high-dimensional DNA methylation data. ML is a subset of artificial intelligence used in data analysis, including algorithms that are trained to automatically recognize data representations, learn from experience, and maximize predictive accuracy (13). A branch of machine learning is deep learning, which is based on artificial neural networks (13). ML methods are capable of analyzing high-dimensional, non-parametric data with complex interactions (13, 14). Although these methods are mainly used for analysis of text and images (13), recent years saw a growing body of epigenome-wide studies that leveraged the power of ML to identify DNA methylation signatures associated with cancer prognosis (Supplementary Figure 1). But thus far no appropriate review has been published to summarize this recent emerging trend. However, the types of ML methods selected and workflows used in different studies are quite heterogeneous, and there might be powerful but yet underused ML methods. Therefore, we conducted a systematic review to comprehensively map studies investigating epigenome-wide associations with cancer prognosis using ML, as well as to identify existing limitations and research gaps.

## Methods

This systematic review was performed and reported according to the Preferred Reporting Items for Systematic Reviews and Meta-Analyses (PRISMA). The complete PRISMA checklist is shown in Supplementary Table 1. The review protocol was published in advance at https://protocolexchange.researchsquare.com/article/pex-1738/v1.

### Search strategy

We searched PubMed, EMBASE, and Web of Science for studies published until 8 June 2022. The search strategy used a combination of Medical Subject Headings (MESH) and key words related to ML, cancer, epigenetic signatures, and prognosis, respectively. The full search strategy is provided in Supplementary Table 2.

Reference lists of included studies were also screened for additional eligible studies. The records retrieved from databases were exported to Endnote 20 (Clarivate Analytics, Philadelphia, USA) and duplicates were removed. Subsequently, the titles and abstracts of the remaining records were screened for eligibility by one author (TY), and potentially eligible studies were retained for full-text review.

### Selection criteria

Studies were deemed eligible if they were peer-reviewed articles, reported in English, included patients with any type of cancer, used an epigenome-wide DNA methylation array, and used at least one ML method to identify DNA methylation signatures (i.e., CpG sites or methylation-driven genes [MDGs]) associated with cancer prognosis (i.e., survival, progression, therapy responses). Multi-omics studies investigating multiple prognostically relevant biomarkers including DNA methylation were also considered. We excluded studies that recruited patients with cancer precursors, used a candidate-gene approach, used ML methods for purposes unrelated to finding prognostically relevant DNA methylation signatures, or simply proposed a customized statistical package or platform based on ML methods.

### Quality assessment

Two authors (TY and ZF) independently assessed the methodological quality of the included studies. The methodological quality of the included studies was assessed by a seven-item checklist adapted from ‘A Tool to Assess Risk of Bias and Applicability of Prediction Model Studies (PROBAST) (15)’ and from the ‘Reporting Recommendations for Tumor Marker Prognostic Studies (REMARK) (16)’.

A study was assigned one point each for meeting the following criteria: total follow-up time >3 years; sample size of training set >100; use of an imputation method for missing data; adjustment for confounders including age, sex, and tumor stage when selecting or evaluating DNA methylation signatures; use of cross-validation or bootstrapping to measure model performance. Additionally, a study could score one point for conducting internal validation, and two points for doing external validation. One point was awarded for measuring the discriminative ability (i.e., the ability of a model to differentiate between high-risk patients and low-risk patients) of selected signatures, and two points in case of additionally measuring calibration accuracy of selected signatures (i.e., the degree of agreement between the predicted and observed prognostic outcomes) of selected signatures.

The quality of each study was rated as low, medium, or high if the score was 0-3, 4-6, or 7-9 points, respectively. Discrepancies between the evaluations of the two authors (TY, ZF) were solved through discussion, and two senior scientists (MH, DE) made the final decision when a consensus was not reached.

### Data extraction and synthesis

The following study-level information was extracted onto pre-designed spreadsheets: first author, year of publication, geographic origin of training set, median follow-up, anatomical site of cancer, cancer stage, prognostic outcome, biospecimen type, methylation array type, methylation signature type, sample size and source of training set, sample size of testing set, type of internal validation, size and source of external validation set, handling of missing values, prognostic model construction, performance measurement, types of ML methods used, and workflows. We only extracted relevant ML methods and workflows that were used to identify individual DNA methylation signatures associated with cancer prognosis, and disregarded methods used for other purposes. The extracted workflows used in each study were cross-checked by a second author (ZF) to ensure accuracy, and disagreements were resolved through discussion between the two authors.

The main study characteristics were summarized in tables, and we summarized the different ML methods by a sunburst chart and a bubble chart, respectively. Workflows used in included studies were summarized by Sankey diagrams. These were performed by using R version 4.1.2 with packages tidyverse, ggplot2, and highcharter (17).

## Results

### Study characteristics

We identified 76 studies that were eligible for this systematic review (Figure 1) (5-9, 18-88).

**Figure 1.**
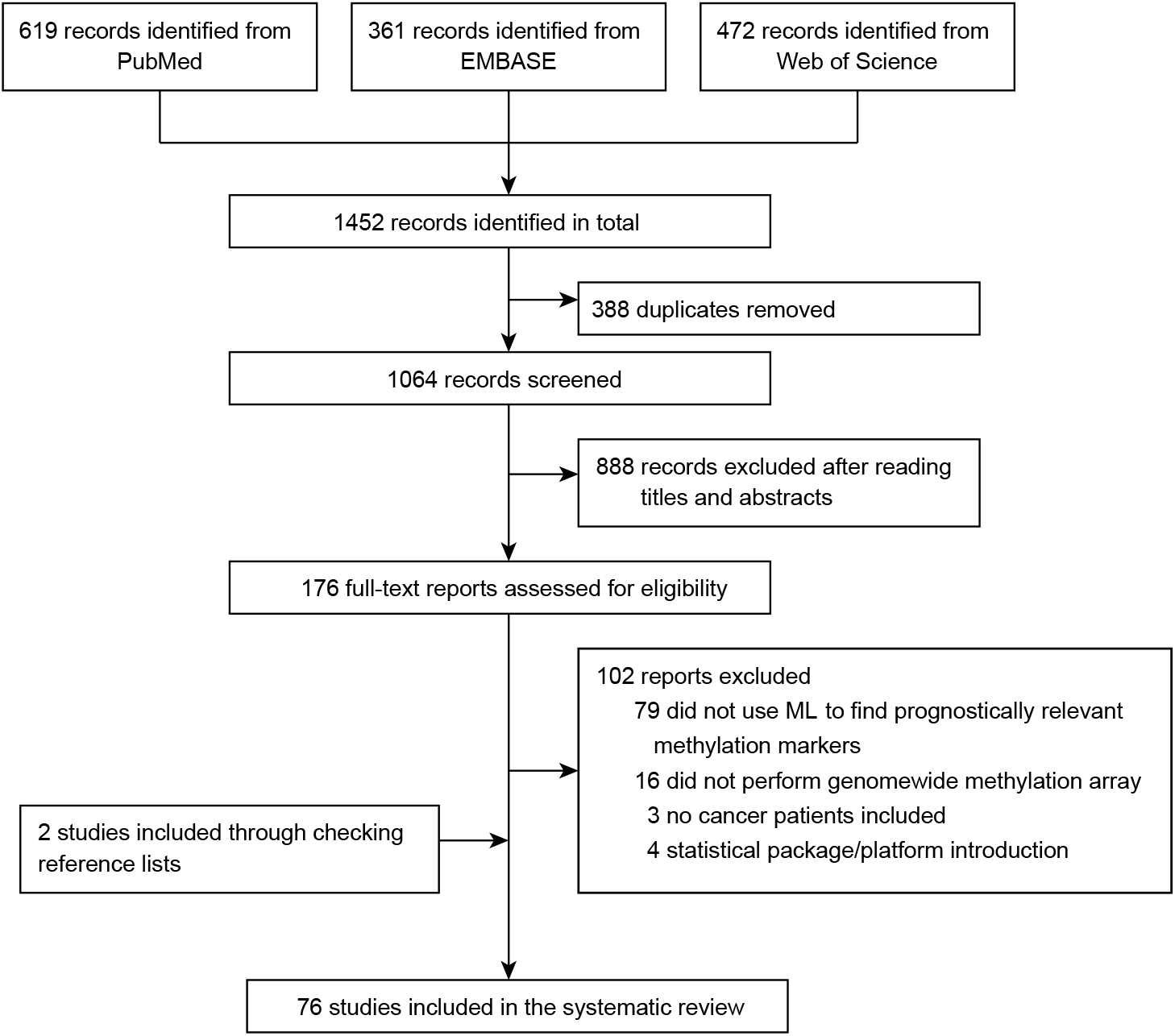
Selection of studies for inclusion. ML = machine learning

Included studies were published between 2002 and 2022, and there was an exponential growth in the number of studies published from 2019 to 2021 (Supplementary Figure 1). Characteristics of included studies are summarized in Table 1, and details of each study are available in Supplementary Table 3.

**Table 1.**
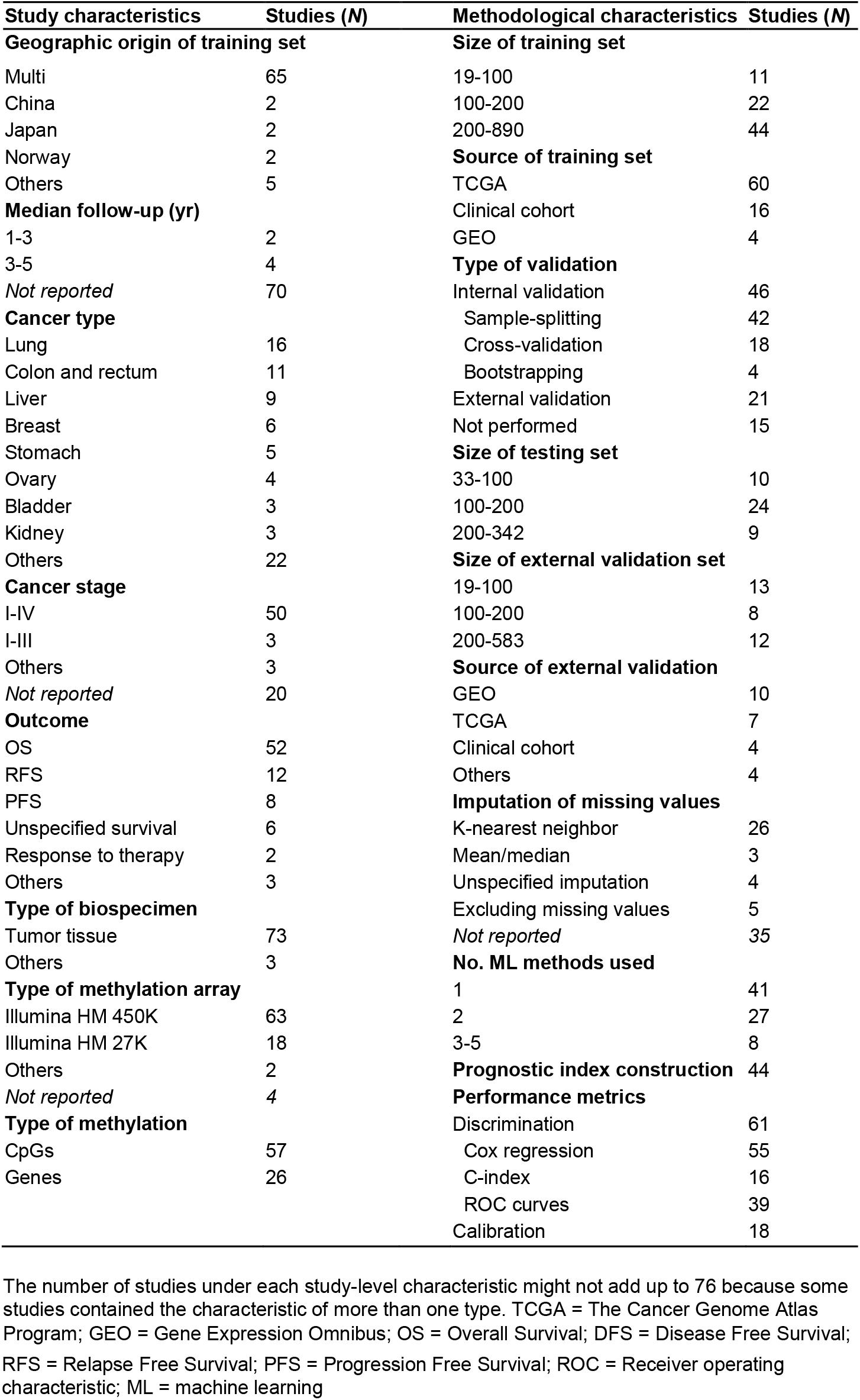
Study and methodological characteristics of included studies.

Lung cancer was the most studied cancer type (*N* = 16), followed by colorectal cancer (*N* = 11). The majority of studies recruited patients with stage I-IV cancer (*N* = 50), and overall survival was the most frequently investigated outcome (*N* = 52). Only six studies based on clinical cohorts reported median follow-up time, ranging from 1.5 to 4.6 years. Nearly all studies determined DNA methylation in tumor tissue (*N* = 73) and used the Illumina 450K BeadChip array (*N* = 63). Fifty-seven studies investigated the role of differentially methylated single CpG sites on cancer prognosis, while 26 studies investigated the prognostic value of MDGs.

In terms of methodological characteristics, more than two thirds of the studies used The Cancer Genome Atlas Program (TCGA) database to identify prognostically relevant DNA methylation biomarkers. The size of the training sets varied from 19 to 890 patients (median: 213). Sample-splitting was the most frequently used internal validation (*N* = 42), and the size of test sets ranged from 33 to 342 patients (median: 143). Twenty-one studies performed external validation, mostly based on the Gene Expression Omnibus (GEO) database. The size of the datasets used for external validation ranged from 12 to 583 (median: 125). Thirty three studies reported imputation of missing values, and K-nearest neighbor was the most frequently used imputation method (*N* = 26). More than half of included studies constructed prognostic models incorporating identified DNA methylation biomarkers.

### Quality assessment

Based on our seven-item quality assessment checklist, 61% of the studies were rated as moderate quality (*N* = 46), and 12 (16%) studies were rated as low quality. The summarized results for each quality assessment item are shown in Figure 2, and the detailed results for each included study are available in Supplementary Table 4.

**Figure 2.**
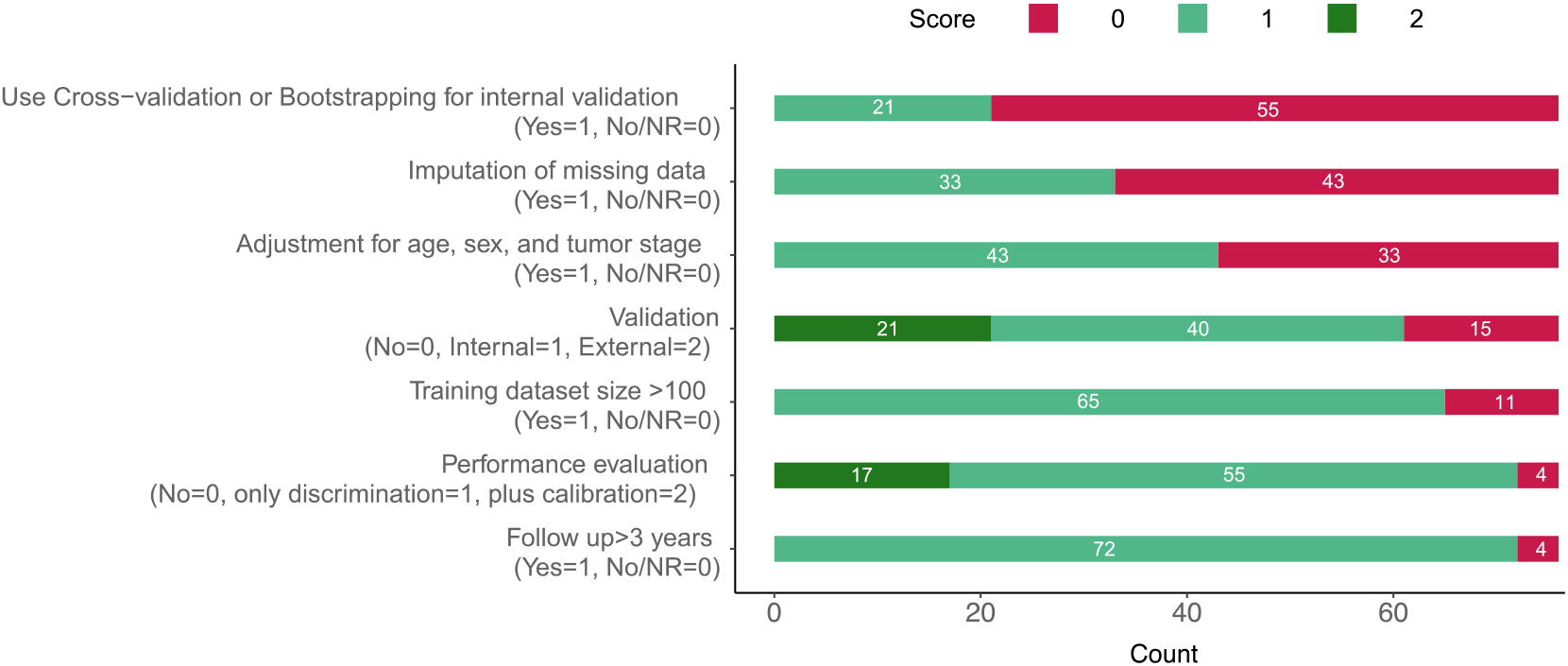
Quality assessment of included studies (N = 69) and distribution of assigned scores. NR = Not reported

The top three items where most studies failed to score points were use of cross-validation or bootstrapping to measure model performance (*N* = 55), use of an imputation method for missing data (*N* = 43), and adjustment for important confounders like age, sex, and tumor stage when selecting or evaluating methylation markers (*N* = 33), respectively.

### ML methods

The complete spectrum of ML methods used in the included studies to identify DNA methylation biomarkers associated with cancer prognosis is shown in Figure 3a. The number of ML methods used by each study ranged from one to five, and the majority of studies used one (*N* = 41) or two (*N* = 27) types of ML methods. The time trend of the frequency of these ML methods used is illustrated in a bubble plot (Figure 3b).

**Figure 3.**
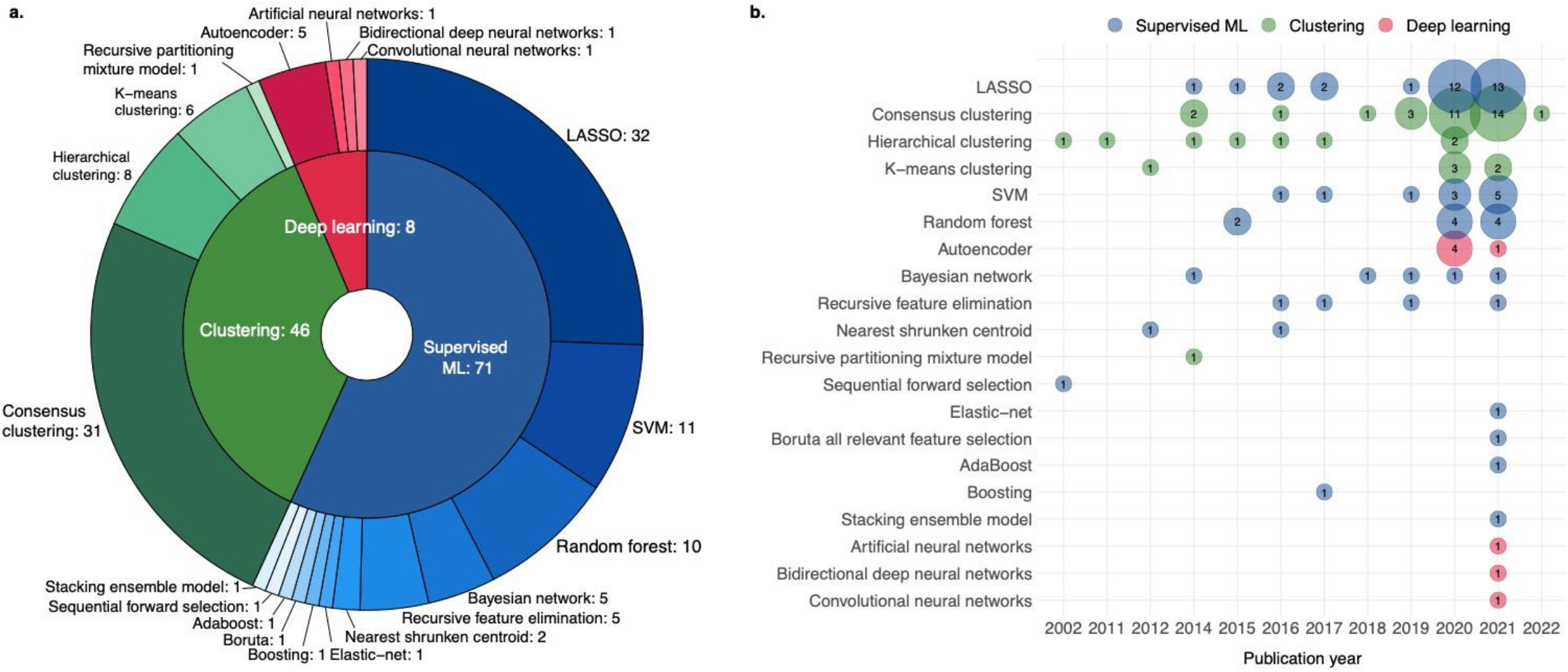
(a) Types and composition of ML methods used in included studies; (b) Frequency of each ML methods used by studies each year. The numbers on the figure do not add up to 76 because they represent the frequency of methods being used, because studies could have used more than one method. In theory, deep learning belongs to machine learning and could be either supervised or unsupervised, but for the purpose of illustration, supervised ML on the figure refers to supervised machine learning algorithms excluding deep learning. The size of a bubble in (b) is proportional to the frequency of the AI method used (the number inside the bubble). ML = machine learning. LASSO = least absolute shrinkage and selection operator.

Supervised machine learning algorithms and unsupervised clustering were predominantly used by a total of 71 and 46 studies, respectively. The top three most frequently used supervised machine learning algorithms were the Least Absolute Shrinkage and Selection Operator (LASSO, *N* = 32), support vector machine (*N* = 11), and random forest (*N* = 10). The most frequently used unsupervised clustering technique was consensus clustering (*N* = 31), followed by hierarchical clustering (*N* = 8) and K-means clustering (*N* = 6). The application of deep learning is a rather recent trend starting from 2020 (Figure 3b), with autoencoder being the most frequently used (*N* = 5).

### Workflows

Based on the characteristic stage, we identified three different workflows: 1) unsupervised clustering, 2) supervised feature selection, and 3) deep learning-based feature transformation. These are visualized in the form of Sankey diagrams (Figure 4). Workflows used by each study are detailed in Supplementary Table 5-7.

**Figure 4.** Workflows used by included studies. At each stage, one study might use more than one method. The number after each method represents the frequency of its being used. ‘Uni’ refers to performing feature selection depending on the *P*-value of univariable analysis, and features with a *P-*value < 0.05 were retained; similarly, ‘Multi’ stands for feature selection based on the *P*-value of multivariable analysis; ‘Cox’ under the stage of model training refers to training a simple multivariable Cox regression model without further feature selection. TT/NT = Tumor tissue/normal tissue. ET/AT = Early-stage tumor/advance-stage tumor. MDG = Methylation driven genes. LASSO = least absolute shrinkage and selection operator. SVM= support vector machine. RFS = recursive feature selection. SVM-RFE = support vector machine-recursive feature elimination. FV-SVM: Forward support vector machine. WGCNA = weighted gene co-expression network analysis. AIC = Akaike information criterion.

**Figure 4.**
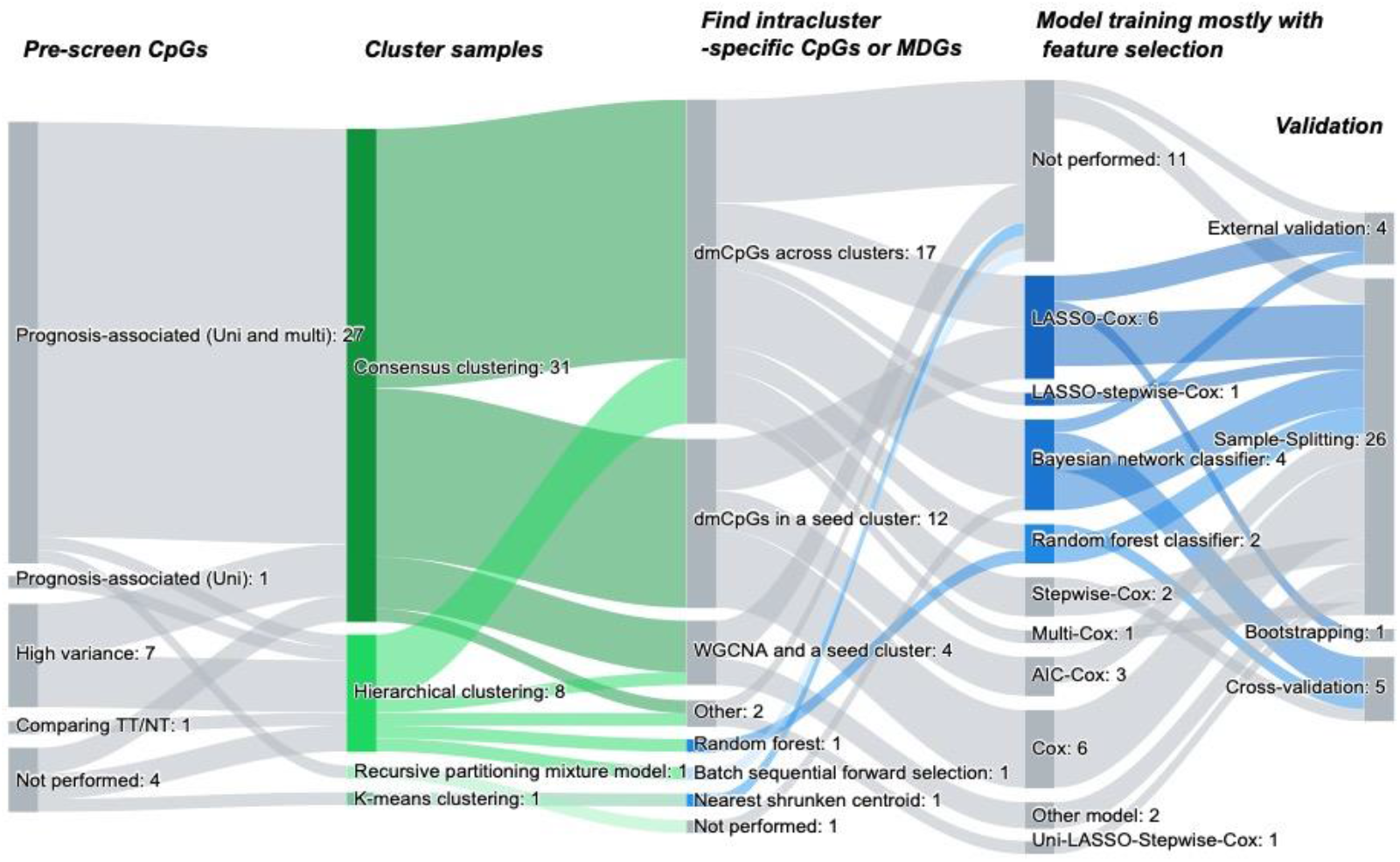
Workflow 1: unsupervised clustering.

**Figure 4.**
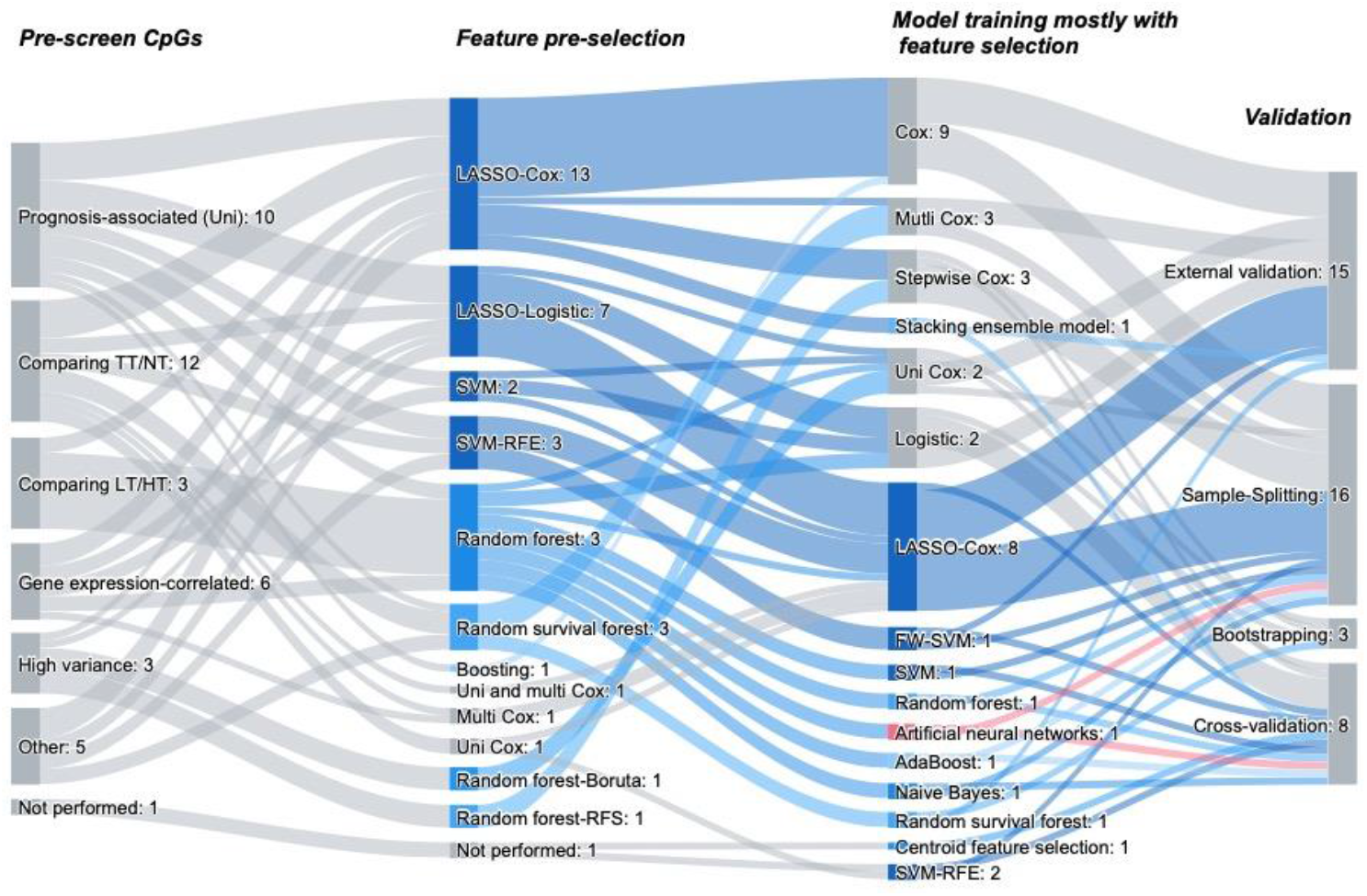
Workflow 2: supervised feature selection.

**Figure 4.**
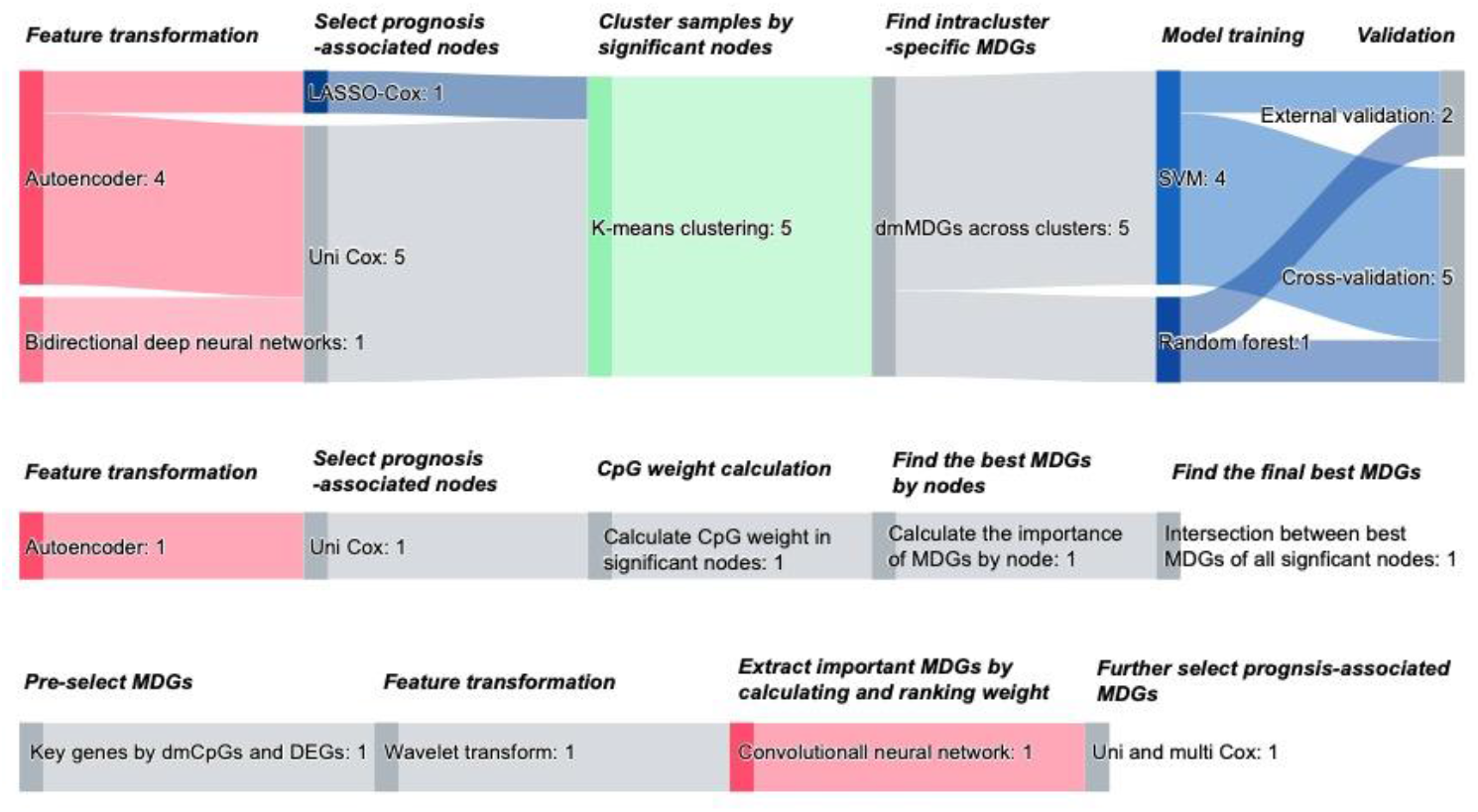
Workflow 3: deep learning-based feature transformation.

#### Workflow 1: unsupervised clustering

Among the 39 studies that adopted clustering-based workflows (5, 18-23, 29, 32-35, 37-39, 43, 45-47, 50, 53-57, 61, 62, 64, 69, 70, 75, 77-80, 83, 84, 87, 88), most started with a preliminary screening of CpG sites (Figure 4.1). The most frequently used pre-selection approach was selecting CpG sites that were significantly associated with prognosis by first performing univariable Cox regression analysis, and then performing multivariable Cox analysis with adjustment for clinical variables. Other pre-screening approaches included the selection of CpG sites above a pre-defined variance threshold, and the selection of significantly differentially methylated CpG sites (dmCpGs) between normal tissue and tumor tissue.

Afterwards, study samples underwent the process of clustering according to their methylation levels of the pre-selected CpG sites, and samples with similar methylation patterns were clustered together. Kaplan-Meier curves and log-rank tests were subsequently used to verify that cancer prognosis across different patient clusters was significantly different. At the third stage, a reduced number of cluster-specific CpG sites were identified, mostly by selecting dmCpG sites across clusters, or by selecting dmCpG sites in the seed cluster (i.e., the cluster associated with good prognosis and containing a great number of dmCpGs). Four studies (50, 56, 62, 79) used weighted gene co-expression network analysis to first identify the co-expression modules having the greatest correlation with the seed cluster, and then to screen for CpGs in that module mostly correlated with the seed cluster.

Based on selected CpGs in the prior stage, eight studies performed additional feature selection, using methods including LASSO (38, 45, 55, 57, 69, 78, 80), stepwise selection (38, 39), and the Akaike information criterion (64, 77), to further refine the number of CpGs. In contrast, four studies (23, 33, 34)(47) directly trained classification models with clusters as labels using Bayesian networks or random forest, while 17 studies constructed prognostic models (37-39, 45, 46, 53-57, 64, 69, 70, 75, 77, 78, 80). Lastly, models developed in the previous stage were mostly internally validated by split-sample validation, albeit four studies validated the model externally in an independent dataset (33, 50, 57, 78).

#### Workflow 2: supervised feature selection

Similar to clustering-based workflows, an initial screening was generally the first step for the 30 studies using supervised feature selection-based workflows (Figure 4.2) (6-9, 24-28, 30, 31, 36, 40-42, 48, 51, 59, 60, 63, 66-68, 71-74, 81, 84, 85). In the next step, nearly all studies used one to three types of supervised feature selection techniques to pre-select prognostically relevant DNA methylation biomarkers prior to training one or more prognostic models. One study (85) used feature selection (random forest) and dimensional reduction (principle component analysis) in parallel before training multiple supervised ML models.

In the stage of model training, further feature selection was performed by more than half of the 30 studies (8, 9, 24, 25, 28, 31, 36, 40-42, 48, 51, 66, 68, 72-74, 81, 84, 85). Sample-splitting was the most commonly used internal model validation method (*N* = 12), and external validation was performed in 15 studies (7, 8, 24-26, 31, 42, 48, 59, 66-68, 72, 74, 81).

As to studies using more than one supervised ML model for feature selection, four studies intersected CpGs selected by different ML techniques (9, 31, 48, 84), whereas two studies only selected the biomarkers retained in one model with the best performance (72, 85). Notably, one study (81) first identified methylation-correlated blocks (MCBs, i.e., blocks of closely co-methylated CpGs) and then selected MCBs associated with survival by LASSO. Next, elastic-net model, support vector regression, and Cox regression were separately trained and used for ranking MCBs. Finally, a stacking ensemble model combining the three models was constructed based on the top-ranked MCB.

#### Workflow 3: deep learning-based feature transformation

We identified a total of seven studies that applied deep learning-based feature transformation to reduce dimensionality (Figure 4.3). In the first step, five studies used autoencoder (44, 49, 52, 58, 76), and one used bidirectional deep neural networks (65), to transform original input features into a reduced number of new features (i.e., hidden nodes), and then they performed univariable Cox regression analysis to select hidden nodes significantly associated with prognosis.

Subsequently, in the five studies (44, 49, 58, 65, 76) that integrated the information of multi-omics biomarkers including DNA methylation, the selected prognosis-associated hidden nodes were used to cluster samples by K-means clustering algorithm. After that, top 20 to 100 significantly differently methylated genes were identified by comparing across clusters. Based on the top features of multi-omics biomarkers, classification models with clusters as labels were constructed by support vector machine or random forest. Cross-validation was used in deep learning-based workflows by five studies, and two studies performed external validation containing DNA methylation information (44, 65).

In contrast, for the one study (52) that exclusively analyzed DNA methylation data, the third step was to calculate the weight of CpGs in every selected hidden node, and then summed the absolute weight values of CpGs corresponding to the same gene. Subsequently, for every significant node, the ratio of weights for every gene was calculated. Lastly, they selected the intersection between the genes with the highest ratio of weights in all the nodes.

In particular, one study (82) applied one-dimensional discrete wavelet transform to pre-selected key genes, and the transformed results were used as the input for the downward convolutional neural network for feature extraction. Afterwards, the top 200 genes were further refined by univariable Cox regression analysis, which were finally used to construct the prognostic model by multivariable Cox regression analysis.

### Performance evaluation

The performance evaluation reported by the included studies varied in terms of objects, metrics, and types of validation (Supplementary Table 8). The objects of evaluation were methylation level-based clusters generated by unsupervised clustering, prognostic models constructed by only incorporating identified DNA methylation biomarkers, and prognostic models based on both DNA methylation markers and clinical variables. The most frequently reported metrics were results of Cox regression analysis (*N* = 55), which were used to evaluate whether there were statistically significant differences in prognosis across clusters, or among patients with different prognostic index. Fewer studies used other metrics, including C-index (*N* =16) and receiver operating characteristic curves (*N* = 39) to evaluate the discriminative power of their prognostic models. Only 18 studies (6, 31, 36, 39-42, 51, 54, 55, 57, 59, 66, 67, 78, 80, 84, 88) evaluated the calibration accuracy of prognostic models.

Given the large heterogeneity across studies in terms of cancer types, prognostic outcomes, methodological strategies, and evaluation metrics, we did not summarize reported metrics. Nevertheless, six studies reported an increase in the discriminative ability of the methylation-based prognostic model after adding other clinical variables (e.g., age, cancer stage) to the model (6, 31, 39, 51, 76, 80).

## Discussion

This systematic review is, to the best of our knowledge, the first review to date that summarizes and catalogs the current use of ML-powered analytic methods in epigenome-wide studies to identify DNA methylation markers associated with cancer prognosis. Following a systematic approach, we reviewed 76 peer-reviewed articles and found pronounced heterogeneity in methodological strategies. Workflows used in prior work were categorized into 1) unsupervised clustering, 2) supervised feature selection, and 3) deep learning-based feature transformation according to the characterized ML techniques used. We also identified several common methodological and reporting issues. This review could potentially serve as a reference for researchers designing ML-based analytic approaches to find meaningful biomarkers, as well as identifying areas warranting further research.

We found that common to all identified workflows, a combination of different dimensionality reduction strategies, with or without the involvement of ML, was applied in more than one stage. These processes are necessary for high-dimensional microarray data, which typically contain many irrelevant and redundant features.

Applying various dimensionality reduction techniques in different stages could improve efficiency by narrowing down the selection scope and reducing computational complexity (89).

Clustering and supervised feature selection were the two main ML-based methods used in the included studies to select a subset of relevant DNA methylation signatures. Consensus clustering, an ensemble-based method of aggregating results from multiple clustering algorithms, was used most frequently, probably because of its relatively robust results compared to other individual clustering algorithms (39).

The most popular supervised feature selection method was LASSO, a regularized regression method that could perform variable selection while constraining overfitting (90). We also identified a recent increase in the use of deep learning methods to compress and extract input features, with autoencoder being the most commonly used.

The popularity of a method is not necessarily equivalent to the optimal or best model selection and performance. An objective evaluation of the efficacy of existing methods and workflows could only be achieved by using the same dataset and benchmarking all approaches against each other through direct comparisons (91). A few identified studies performed comparative analysis of the autoencoder framework with other methods (49, 52, 65, 76), and results mostly favored the autoencoder approach. Besides, one study showed that wavelet-based deep learning model outperform traditional LASSO and other wavelet-based approaches (82). However, we did not find benchmarking studies comparing the three types of workflows identified by this review.

For supervised feature selection and unsupervised approaches, each has its own advantages and disadvantages. Specifically, supervised learning uses labeled data in which the targeted variable to be predicted is known. However, it is costly and time-consuming to follow up all patients in large cohorts or databases for a reasonably long time to obtain information about the targeted outcome (89). In comparison, unsupervised learning does not require outcome information and may thus enable analyses in much larger patient cohorts. But it has the downside that the results are not necessarily related to a meaningful clinical outcome (89). As a remedy semi-supervised approaches might be used, which could combine the best of both approaches. Alternatively, studies might use unsupervised and supervised approaches in conjunction at different stages to select features, as done in some of the studies (38, 45, 55, 57, 69, 78, 80).

Many methodological and reporting problems were common in the identified studies. First, only five out of the 69 included studies reported median follow-up time (6, 24, 26, 29, 31), which is important information because survival outcomes can only be decently evaluated after several years of follow-up, especially if patients with early stages of cancer are included. None of the studies using TCGA data reported median follow-up time, so little can be said about the data quality and interpretability of the conducted study. Second, six studies failed to specify the type of survival outcomes (e.g., overall survival or disease-free survival) (43, 46, 47, 60, 62, 75), which further limits the interpretation of results.

Third, handling of missing values was not specified in nearly half of the studies, in which patients with any missing data were likely to be omitted. Six studies explicitly reported exclusion of patients with missing values (6, 44, 46, 52, 57, 76). Simply excluding patients with any missing data from the analysis could lead to selection bias, and proper imputation methods ought to be in place (15, 92).

Fourth, the association between aberrant DNA methylation signatures and cancer prognosis is only relevant when they provide additional prognostic value not captured by known prognostic factors such as tumor stage, age and sex. But more than half of the identified studies did not take these important clinical factors into account.

Fifth, none of the studies mentioned how competing risk events were dealt with in the analysis when the outcome of interest was not overall survival. Sixth, most studies only used one single supervised learning model for variable selection without providing a rationale why choosing that specific algorithm. The reasons for selecting a modeling method should be clearly stated (92). There might have been room for improvement in model performance if a wider range of models had been tried and compared in these studies.

Seventh, the aim of internal validation is to evaluate the performance of a model in a dataset not involved in model training (92). However, one study randomly selected a subset from the training set to validate the model (63), which is not a real internal validation. Such an approach could lead to data leakage and thereby to an optimistically biased evaluation (92).

Most studies randomly split a single dataset into training and test sets. This approach fails to efficiently use all data available for model training, and the model performance can change every run when the dataset is randomly split in different ways (15, 92).

Better internal validation methods such as cross-validation and bootstrapping are recommended by guidelines(15, 92). It is also recommended that all analysis steps (imputation, screening etc.) should be incorporated in the cross-validation procedures to prevent data leakage (92). Moreover, external validation is crucial to verify the generalizability of findings, which is especially important for high-dimensional data with high risk of overfitting.

Eighth, discrimination and calibration are two important metrics to evaluate the performance of prognostic models incorporating meaningful DNA methylation biomarkers (15, 92). However, most studies reviewed focused on discrimination metrics, whereas calibration accuracy was only measured by less than one quarter of included studies. Both discrimination and calibration evaluation metrics are required for all datasets including training, testing, and external validation sets(92). However, 5 studies reported discrimination metrics in both the testing set and external validation set, but reported calibration only in the training set (42, 48, 67, 68, 78).

Lastly, we found that 18 identified studies first constructed a Cox prognostic model based on single selected CpG sites (i.e., methylation score), which was subsequently used as a single predictor, together with other clinical variables, to construct a second Cox prognostic model (6, 31, 36, 39-42, 55, 57, 66-69, 73, 78, 80, 81). It might be an inappropriate approach to construct the first “methylation score”, in which the relative weight of individual CpGs were consequently fixed for the second prognostic model. Moreover, four studies dichotomized the continuous methylation score (31, 41, 55, 59), leading to loss of information (15). Given that clinical information and DNA methylation biomarkers are different types of information characterized by different statistical properties, they could be combined by multimodal deep learning technique. Additionally, traditional nomograms were used by all the 18 studies for the graphical presentation of their prediction model. More user-friendly online risk calculator platforms are recommended over nomograms in today’s digital era (92, 93).

## Limitation of the study

This systematic review is the first of its kind and has some limitations. First, we specifically included only studies using a genome-wide array to identify DNA methylation signatures associated with cancer prognosis. We recognize that similar ML approaches can be applied to non-genome-wide studies or to the identification of other biomarkers (e.g., mRNA) relevant to other diseases. Second, in the included multi-omics studies (44, 49, 58, 65, 76), identifying individual prognostically relevant DNA methylation biomarkers were merely side products instead of primary study aims. Third, variations in disease outcomes and the lack of benchmarking studies prevented us from comparing the performance of identified workflows.

Further benchmarking studies are needed to systematically evaluate the comparative performance of various ML-based methods and workflows identified in this review and to identify the best approaches. Novel ML methods not included in this review may also help to reduce dimensionality and to find prognostically relevant biomarkers. Future epigenome-wide association studies intending to use ML methods should carefully adhere to relevant methodological and reporting standards (15, 92, 94), so as to improve methodological quality and reproducibility.

## Conclusions

wide association studies. We identified three major categories of ML-based workflows as well as some common methodological and reporting flaws in existing studies. Benchmarking studies are needed to compare the relative performance of these workflows using a same dataset. Adherence to methodological and reporting guidelines is strongly recommended for future research in this area.

## Data Availability

All data produced in the present work are contained in the manuscript and supplement.

## Abbreviations

ML: machine learning;
LASSO: least absolute shrinkage and selection operator;
TNM: tumor-lymph node-metastasis;
PRISMA: Preferred Reporting Items for Systematic reviews and Meta-Analyses;
MESH: Medical Subject Headings;
MDGs: methylation-driven genes;
PROBAST: A Tool to Assess Risk of Bias and Applicability of Prediction Model Studies;
RMARK: Reporting Recommendations for Tumor Marker Prognostic Studies;
TCGA: The Cancer Genome Atlas Program;
GEO: Gene Expression Omnibus;
dmCpGs: differentially methylated CpG sites;

## Ethics approval and consent to participate

Not applicable.

## Consent for publication

Not applicable.

## Availability of data and materials

The datasets supporting the conclusions of this article are included within the supplementary materials of the article.

## Authors’ contribution

MH conceived the study. MH and TY designed the protocol. DE provided statistical input and consultation. TY conducted study selection, and TY and ZF conducted data extraction and quality assessment. TY made tables and figures and drafted the manuscript. DE contributed to interpretation of findings. DE, ZF, EA, and JNK, and HB critically revised the manuscript. MH supervised the study.

## Acknowledgements

We thank Xinyi Zhou for helping retrieve initial records from the EMBASE database.

## References

1. Sung H, Ferlay J, Siegel RL, Laversanne M, Soerjomataram I, Jemal A, et al. Global Cancer Statistics 2020: GLOBOCAN Estimates of Incidence and Mortality Worldwide for 36 Cancers in 185 Countries. CA Cancer J Clin. 2021;71(3):209–49.

2. Gospodarowicz M, Brierley J, Wittekind C. TNM Classification of Malignant Tumours, 8th Edition. UK: Wiley-Blackwell; 2017.

3. Takahashi S, Asada K, Takasawa K, Shimoyama R, Sakai A, Bolatkan A, et al. Predicting Deep Learning Based Multi-Omics Parallel Integration Survival Subtypes in Lung Cancer Using Reverse Phase Protein Array Data. Biomolecules. 2020;10(10):1460.

4. Xue W, Wu X, Wang F, Han P, Cui B. Genome-wide methylation analysis identifies novel prognostic methylation markers in colon adenocarcinoma. Biomed Pharmacother. 2018;108:288–96.

5. Ouchi K, Takahashi S, Yamada Y, Tsuji S, Tatsuno K, Takahashi H, et al. DNA methylation status as a biomarker of anti-epidermal growth factor receptor treatment for metastatic colorectal cancer. Cancer Sci. 2015;106(12):1722–9.

6. Luo H, Zhao Q, Wei W, Zheng L, Yi S, Li G, et al. Circulating tumor DNA methylation profiles enable early diagnosis, prognosis prediction, and screening for colorectal cancer. Sci Transl Med. 2020;12(524):eaax7533.

7. Fleischer T, Frigessi A, Johnson KC, Edvardsen H, Touleimat N, Klajic J, et al. Genome-wide DNA methylation profiles in progression to in situ and invasive carcinoma of the breast with impact on gene transcription and prognosis. Genome Biol. 2014;15(8):435.

8. Dong RZ, Yang X, Zhang XY, Gao PT, Ke AW, Sun HC, et al. Predicting overall survival of patients with hepatocellular carcinoma using a three-category method based on DNA methylation and machine learning. J Cell Mol Med. 2019;23(5):3369–74.

9. Bedon L, Dal Bo M, Mossenta M, Busato D, Toffoli G, Polano M. A Novel Epigenetic Machine Learning Model to Define Risk of Progression for Hepatocellular Carcinoma Patients. Int J Mol Sci. 2021;22(3):1075.

10. Harrison A, Parle-McDermott A. DNA methylation: a timeline of methods and applications. Front Genet. 2011;2:74.

11. Yong W-S, Hsu F-M, Chen P-Y. Profiling genome-wide DNA methylation. Epigenetics & Chromatin. 2016;9(1):26.

12. Gareth J, Daniela W, Trevor H, Robert T. An Introduction to Statistical Learning: with Applications in R (Second Edition). New York: Springer; 2021. 261–7 p.

13. Nichols JA, Herbert Chan HW, Baker MAB. Machine learning: applications of artificial intelligence to imaging and diagnosis. Biophys Rev. 2019;11(1):111–8.

14. Bostwick DG, Burke HB. Prediction of individual patient outcome in cancer: comparison of artificial neural networks and Kaplan--Meier methods. Cancer. 2001;91(8 Suppl):1643–6.

15. Moons KGM, Wolff RF, Riley RD, Whiting PF, Westwood M, Collins GS, et al. PROBAST: A Tool to Assess Risk of Bias and Applicability of Prediction Model Studies: Explanation and Elaboration. Ann Intern Med. 2019;170(1):W1–W33.

16. Sauerbrei W, Taube SE, McShane LM, Cavenagh MM, Altman DG. Reporting Recommendations for Tumor Marker Prognostic Studies (REMARK): An Abridged Explanation and Elaboration. J Natl Cancer Inst. 2018;110(8):803–11.

17. R Core Team (2021). R: A language and environment for statistical computing. R Foundation for Statistical Computing, Vienna, Austria. [Available from: https://www.R-project.org/.

18. Wei SH, Chen CM, Strathdee G, Harnsomburana J, Shyu CR, Rahmatpanah F, et al. Methylation microarray analysis of late-stage ovarian carcinomas distinguishes progression-free survival in patients and identifies candidate epigenetic markers. Clin Cancer Res. 2002;8(7):2246–52.

19. Walker BA, Wardell CP, Chiecchio L, Smith EM, Boyd KD, Neri A, et al. Aberrant global methylation patterns affect the molecular pathogenesis and prognosis of multiple myeloma. Blood. 2011;117(2):553–62.

20. Sigalotti L, Covre A, Fratta E, Parisi G, Sonego P, Colizzi F, et al. Whole genome methylation profiles as independent markers of survival in stage IIIC melanoma patients. J Transl Med. 2012;10:185.

21. Chambwe N, Kormaksson M, Geng H, De S, Michor F, Johnson NA, et al. Variability in DNA methylation defines novel epigenetic subgroups of DLBCL associated with different clinical outcomes. Blood. 2014;123(11):1699–708.

22. Mah WC, Thurnherr T, Chow PK, Chung AY, Ooi LL, Toh HC, et al. Methylation profiles reveal distinct subgroup of hepatocellular carcinoma patients with poor prognosis. PLoS One. 2014;9(8):e104158.

23. Wang C, Cicek MS, Charbonneau B, Kalli KR, Armasu SM, Larson MC, et al. Tumor hypomethylation at 6p21.3 associates with longer time to recurrence of high-grade serous epithelial ovarian cancer. Cancer Res. 2014;74(11):3084–91.

24. Villanueva A, Portela A, Sayols S, Battiston C, Hoshida Y, Méndez-González J, et al. DNA methylation-based prognosis and epidrivers in hepatocellular carcinoma. Hepatology. 2015;61(6):1945–56.

25. Wei JH, Haddad A, Wu KJ, Zhao HW, Kapur P, Zhang ZL, et al. A CpG-methylation-based assay to predict survival in clear cell renal cell carcinoma. Nat Commun. 2015;6:8699.

26. Bjaanaes MM, Fleischer T, Halvorsen AR, Daunay A, Busato F, Solberg S, et al. Genome-wide DNA methylation analyses in lung adenocarcinomas: Association with EGFR, KRAS and TP53 mutation status, gene expression and prognosis. Mol Oncol. 2016;10(2):330–43.

27. Lim AM, Wong NC, Pidsley R, Zotenko E, Corry J, Dobrovic A, et al. Genome-scale methylation assessment did not identify prognostic biomarkers in oral tongue carcinomas. Clin Epigenetics. 2016;8(1):74.

28. Lu J, Cowperthwaite MC, Burnett MG, Shpak M. Molecular Predictors of Long-Term Survival in Glioblastoma Multiforme Patients. PLoS One. 2016;11(4):e0154313.

29. Saito Y, Nagae G, Motoi N, Miyauchi E, Ninomiya H, Uehara H, et al. Prognostic significance of CpG island methylator phenotype in surgically resected small cell lung carcinoma. Cancer Sci. 2016;107(3):320–5.

30. Hao X, Luo H, Krawczyk M, Wei W, Wang W, Wang J, et al. DNA methylation markers for diagnosis and prognosis of common cancers. Proc Natl Acad Sci U S A. 2017;114(28):7414–9.

31. Qiu J, Peng B, Tang Y, Qian Y, Guo P, Li M, et al. CpG Methylation Signature Predicts Recurrence in Early-Stage Hepatocellular Carcinoma: Results From a Multicenter Study. J Clin Oncol. 2017;35(7):734–42.

32. Stieglitz E, Mazor T, Olshen AB, Geng H, Gelston LC, Akutagawa J, et al. Genome-wide DNA methylation is predictive of outcome in juvenile myelomonocytic leukemia. Nat Commun. 2017;8(1):2127.

33. Zhang S, Wang Y, Gu Y, Zhu J, Ci C, Guo Z, et al. Specific breast cancer prognosis-subtype distinctions based on DNA methylation patterns. Mol Oncol. 2018;12(7):1047–60.

34. Chen WB, Zhuang J, Wang PP, Jiang JJ, Lin CH, Zeng P, et al. DNA methylation-based classification and identification of renal cell carcinoma prognosis-subgroups. Cancer Cell Int. 2019;19:185.

35. Li X, Cai Y. Better prognostic determination and feature characterization of cutaneous melanoma through integrative genomic analysis. Aging (Albany NY). 2019;11(14):5081–107.

36. Wang Y, Deng H, Xin S, Zhang K, Shi R, Bao X. Prognostic and predictive value of three DNA methylation signatures in lung adenocarcinoma. Front Genet. 2019;10:349.

37. Yang C, Zhang Y, Xu X, Li W. Molecular subtypes based on DNA methylation predict prognosis in colon adenocarcinoma patients. Aging (Albany NY). 2019;11(24):11880–92.

38. Bian J, Long JY, Yang X, Yang XB, Xu YY, Lu X, et al. Signature based on molecular subtypes of deoxyribonucleic acid methylation predicts overall survival in gastric cancer. World J Gastroenterol. 2020;26(41):6414–30.

39. Cai Q, He B, Xie H, Zhang P, Peng X, Zhang Y, et al. Identification of a novel prognostic DNA methylation signature for lung adenocarcinoma based on consensus clustering method. Cancer Med. 2020;9(20):7488–502.

40. Chen H, Ma X, Yang M, Wang M, Li L, Huang T. A methylomics-associated nomogram predicts recurrence-free survival of thyroid papillary carcinoma. Cancer Med. 2020;9(19):7183–93.

41. Deng Y, Wan H, Tian J, Cheng X, Rao M, Li J, et al. CpG-methylation-based risk score predicts progression in colorectal cancer. Epigenomics. 2020;12(7):605–15.

42. Dong M, Yang Z, Li X, Zhang Z, Yin A. Screening of methylation gene sites as prognostic signature in lung adenocarcinoma. Yonsei Med J. 2020;61(12):1013–23.

43. He H, Chen D, Cui SM, Wu G, Piao HL, Wang X, et al. HDNA methylation data-based molecular subtype classification related to the prognosis of patients with hepatocellular carcinoma. BMC Med Genomics. 2020;13(1):118.

44. Lee TY, Huang KY, Chuang CH, Lee CY, Chang TH. Incorporating deep learning and multi-omics autoencoding for analysis of lung adenocarcinoma prognostication. Comput Biol Chem. 2020;87:107277.

45. Li C, Ke J, Liu J, Su J. DNA methylation data-based molecular subtype classification related to the prognosis of patients with cervical cancer. J Cell Biochem. 2020;121(3):2713–24.

46. Li TD, Chen X, Gu ML, Deng AM, Qian C. Identification of the subtypes of gastric cancer based on DNA methylation and the prediction of prognosis. Clin Epigenetics. 2020;12(1):161.

47. Lian QX, Wang B, Fan LJ, Sun JQ, Wang GL, Zhang JD. DNA methylation data-based molecular subtype classification and prediction in patients with gastric cancer. Cancer Cell Int. 2020;20(1):349.

48. Luo R, Song J, Xiao X, Xie Z, Zhao Z, Zhang W, et al. Identifying CpG methylation signature as a promising biomarker for recurrence and immunotherapy in non-small-cell lung carcinoma. Aging (Albany NY). 2020;12(14):14649–76.

49. Lv J, Wang J, Shang X, Liu F, Guo S. Survival prediction in patients with colon adenocarcinoma via multiomics data integration using a deep learning algorithm. Biosci Rep. 2020;40(12):BSR20201482.

50. Ma H, Zhao C, Zhao Z, Hu L, Ye F, Wang H, et al. Specific glioblastoma multiforme prognostic-subtype distinctions based on DNA methylation patterns. Cancer Gene Ther. 2020;27(9):702–14.

51. Ma X, Chen H, Wang G, Li L, Tao K. DNA methylation profiling to predict overall survival risk in gastric cancer: development and validation of a nomogram to optimize clinical management. J Cancer. 2020;11(15):4352–65.

52. Macías-García L, Martínez-Ballesteros M, Luna-Romera JM, García-Heredia JM, García-Gutiérrez J, Riquelme-Santos JC. Autoencoded DNA methylation data to predict breast cancer recurrence: Machine learning models and gene-weight significance. Artif Intell Med. 2020;110:101976.

53. Shi S, Xu M, Xi Y. Molecular subtypes based on DNA promoter methylation predict prognosis in lung adenocarcinoma patients. Aging (Albany NY). 2020;12(23):23917–30.

54. Tian ZJ, Meng LF, Long XB, Diao TX, Hu ML, Wang M, et al. DNA methylation-based classification and identification of bladder cancer prognosis-associated subgroups. Cancer Cell Int. 2020;20(1):255.

55. Wang X, Wang D, Liu J, Feng M, Wu X. A novel CpG-methylation-based nomogram predicts survival in colorectal cancer. Epigenetics. 2020;15(11):1213–27.

56. Wang Y, Wang Y, Wang Y, Zhang Y. Identification of prognostic signature of non-small cell lung cancer based on TCGA methylation data. Sci Rep. 2020;10(1):8575.

57. Xiang R, Fu T. Gastrointestinal adenocarcinoma analysis identifies promoter methylation-based cancer subtypes and signatures. Sci Rep. 2020;10(1):21234.

58. Yu J, Wu X, Lv M, Zhang Y, Zhang X, Li J, et al. A model for predicting prognosis in patients with esophageal squamous cell carcinoma based on joint representation learning. Oncol Lett. 2020;20(6):387.

59. Zhang C, Wang F, Guo F, Ye C, Yang Y, Huang Y, et al. A 13-gene risk score system and a nomogram survival model for predicting the prognosis of clear cell renal cell carcinoma. Urol Oncol. 2020;38(3):74.e1-.e11.

60. Zhang E, Hou X, Hou B, Zhang M, Song Y. A risk prediction model of DNA methylation improves prognosis evaluation and indicates gene targets in prostate cancer. Epigenomics. 2020;12(4):333–52.

61. Zheng J, Zhang T, Guo W, Zhou C, Cui X, Gao L, et al. Integrative Analysis of Multi-Omics Identified the Prognostic Biomarkers in Acute Myelogenous Leukemia. Front Oncol. 2020;10:591937.

62. Chen H, Qin Q, Xu Z, Chen T, Yao X, Xu B, et al. DNA methylation data-based prognosis-subtype distinctions in patients with esophageal carcinoma by bioinformatic studies. J Cell Physiol. 2021;236(3):2126–38.

63. Hao XY, Li AQ, Shi H, Guo TK, Shen YF, Deng Y, et al. A novel DNA methylation-based model that effectively predicts prognosis in hepatocellular carcinoma. Biosci Rep. 2021;41(3):BSR20203945.

64. Huang G, Zhang J, Gong L, Liu D, Wang X, Chen Y, et al. Specific Lung Squamous Cell Carcinoma Prognosis-Subtype Distinctions Based on DNA Methylation Patterns. Med Sci Monit. 2021;27:e929524.

65. Huang GJ, Wang C, Fu X. Bidirectional deep neural networks to integrate RNA and DNA data for predicting outcome for patients with hepatocellular carcinoma. Future Oncol. 2021;17(33):4481–95.

66. Huang H, Fu JM, Zhang L, Xu J, Li DP, Onwuka JU, et al. Integrative Analysis of Identifying Methylation-Driven Genes Signature Predicts Prognosis in Colorectal Carcinoma. Front Oncol. 2021;11:629860.

67. Huang W, Weng W, Wu B, Ye T, Lin Z, Zhang Z, et al. Development and validation of the trans-omics model for pancreatic adenocarcinoma. Epigenomics. 2021;13(1):15–30.

68. Lee N, Xia X, Meng H, Zhu W, Wang X, Zhang T, et al. Identification of a novel CpG methylation signature to predict prognosis in lung squamous cell carcinoma. Cancer Biomarkers. 2021;30(1):63–73.

69. Li K, Wu Z, Yao J, Fan J, Wei Q. DNA methylation patterns-based subtype distinction and identification of soft tissue sarcoma prognosis. Medicine (Baltimore). 2021;100(5):e23787.

70. Li XS, Nie KC, Zheng ZH, Zhou RS, Huang YS, Ye ZJ, et al. Molecular subtypes based on DNA methylation predict prognosis in lung squamous cell carcinoma. BMC Cancer. 2021;21(1):96.

71. Peng YJ, Zhao J, Yin F, Sharen G, Wu QY, Chen Q, et al. A methylation-driven gene panel predicts survival in patients with colon cancer. Febs Open Bio. 2021;11(9):2490–506.

72. Shang SP, Li X, Gao Y, Guo S, Sun DL, Zhou HX, et al. MeImmS: Predict Clinical Benefit of Anti-PD-1/PD-L1 Treatments Based on DNA Methylation in Non-small Cell Lung Cancer. Front Genet. 2021;12:676449.

73. Sun GP, Duan H, Xing YH, Zhang DW. Prognostic Score Model Based on Ten Differentially Methylated Genes for Predicting Clinical Outcomes in Patients with Adenocarcinoma of the Colon. Cancer Manag Res. 2021;13:5113–25.

74. Tan S, Gui WW, Wang SM, Sun CQ, Xu X, Liu LX. A methylation-based prognostic model predicts survival in patients with colorectal cancer. J Gastrointest Oncol. 2021;12(4):1590–600.

75. Wang L, Xue FX. Prognosis-associated methylation subtypes in endometrial carcinoma patients and the role of magnetic nanoparticles in gene extraction. Materials Express. 2021;11(8):1288–98.

76. Wang TH, Lee CY, Lee TY, Huang HD, Hsu JBK, Chang TH. Biomarker Identification through Multiomics Data Analysis of Prostate Cancer Prognostication Using a Deep Learning Model and Similarity Network Fusion. Cancers (Basel). 2021;13(11):2528.

77. Wu ZH, Tang Y, Zhou Y. DNA Methylation Based Molecular Subtypes Predict Prognosis in Breast Cancer Patients. Cancer Control. 2021;28:1073274820988519.

78. Xu QH, Hu YB, Chen SH, Zhu YL, Li SW, Shen F, et al. Immunological Significance of Prognostic DNA Methylation Sites in Hepatocellular Carcinoma. Front Mol Biosci. 2021;8:683240.

79. Yin LL, Zhang NN, Yang Q. DNA methylation subtypes for ovarian cancer prognosis. Febs Open Bio. 2021;11(3):851–65.

80. Yin XL, Kong LM, Liu P. Identification of prognosis-related molecular subgroups based on DNA methylation in pancreatic cancer. Clin Epigenetics. 2021;13(1):109.

81. Yu X, Yang Q, Wang D, Li Z, Chen N, Kong DX. Predicting lung adenocarcinoma disease progression using methylation-correlated blocks and ensemble machine learning classifiers. PeerJ. 2021;9.

82. Tang B, Chen Y, Wang Y, Nie J. A Wavelet-Based Learning Model Enhances Molecular Prognosis in Pancreatic Adenocarcinoma. Biomed Res Int. 2021;2021:7865856.

83. Zhu CM, Zhang SY, Liu D, Wang QQ, Yang NN, Zheng ZW, et al. A Novel Gene Prognostic Signature Based on Differential DNA Methylation in Breast Cancer. Frontiers in Genetics. 2021;12.

84. Guo YF, Yin JJ, Dai YH, Guan YD, Chen PJ, Chen YQ, et al. A Novel CpG Methylation Risk Indicator for Predicting Prognosis in Bladder Cancer. Front Cell Dev Biol. 2021;9.

85. Kutlay A, Son YA. Integrative Predictive Modeling of Metastasis in Melanoma Cancer Based on MicroRNA, mRNA, and DNA Methylation Data. Frontiers in Molecular Biosciences. 2021;8.

86. Wang JY, Li J, Chen RF, Yue HR, Li WZ, Wu BB, et al. DNA methylation-based profiling reveals distinct clusters with survival heterogeneity in high-grade serous ovarian cancer. Clinical Epigenetics. 2021;13(1).

87. Xu DD, Li C, Zhang YJ, Zhang JZ. DNA methylation molecular subtypes for prognosis prediction in lung adenocarcinoma. BMC Pulm Med. 2022;22(1).

88. Zhou Q, Chen QY, Chen X, Hao L. Bioinformatics analysis to screen DNA methylation-driven genes for prognosis of patients with bladder cancer. Transl Androl Urol. 2021;10(9):3604-+.

89. Ang JC, Mirzal A, Haron H, Hamed HN. Supervised, Unsupervised, and Semi-Supervised Feature Selection: A Review on Gene Selection. IEEE/ACM Trans Comput Biol Bioinform. 2016;13(5):971–89.

90. Tibshirani R. The lasso method for variable selection in the Cox model. Stat Med. 1997;16(4):385–95.

91. Morid MA, Borjali A, Del Fiol G. A scoping review of transfer learning research on medical image analysis using ImageNet. Comput Biol Med. 2021;128:104115.

92. de Hond AAH, Leeuwenberg AM, Hooft L, Kant IMJ, Nijman SWJ, van Os HJA, et al. Guidelines and quality criteria for artificial intelligence-based prediction models in healthcare: a scoping review. NPJ Digit Med. 2022;5(1):2.

93. Park SY. Nomogram: An analogue tool to deliver digital knowledge. J Thorac Cardiovasc Surg. 2018;155(4):1793.

94. Moons KG, Altman DG, Reitsma JB, Ioannidis JP, Macaskill P, Steyerberg EW, et al. Transparent Reporting of a multivariable prediction model for Individual Prognosis or Diagnosis (TRIPOD): explanation and elaboration. Ann Intern Med. 2015;162(1):W1–73.

