## Supplement for "Machine learning in the identification of prognostic DNA methylation biomarkers among patients with cancer: a systematic review of epigenome-wide studies"

**Abbreviations used in this supplement**

ML = machine learning
PRISMA = Preferred Reporting Items for Systematic reviews and Meta-Analyses
NR = Not reported
PROBAST = A Tool to Assess Risk of Bias and Applicability of Prediction Model Studies
RMARK = Reporting Recommendations for Tumor Marker Prognostic Studies
TCGA = The Cancer Genome Atlas Program
GEO = Gene Expression Omnibus
OS = Overall Survival
DFS = Disease Free Survival
RFS = Relapse Free Survival
PFS = Progression Free Survival
PI = Prognostic index
ROC = Receiver operating characteristic
AUC = Area under curves
LASSO = least absolute shrinkage and selection operator
SVM-RFE = support vector machine-recursive feature elimination
FV-SVM: Forward support vector machine
WGCNA = weighted gene co-expression network analysis
AIC = Akaike information criterion
TT/NT = Tumor tissue/normal tissue
MDG = Methylation driven genes

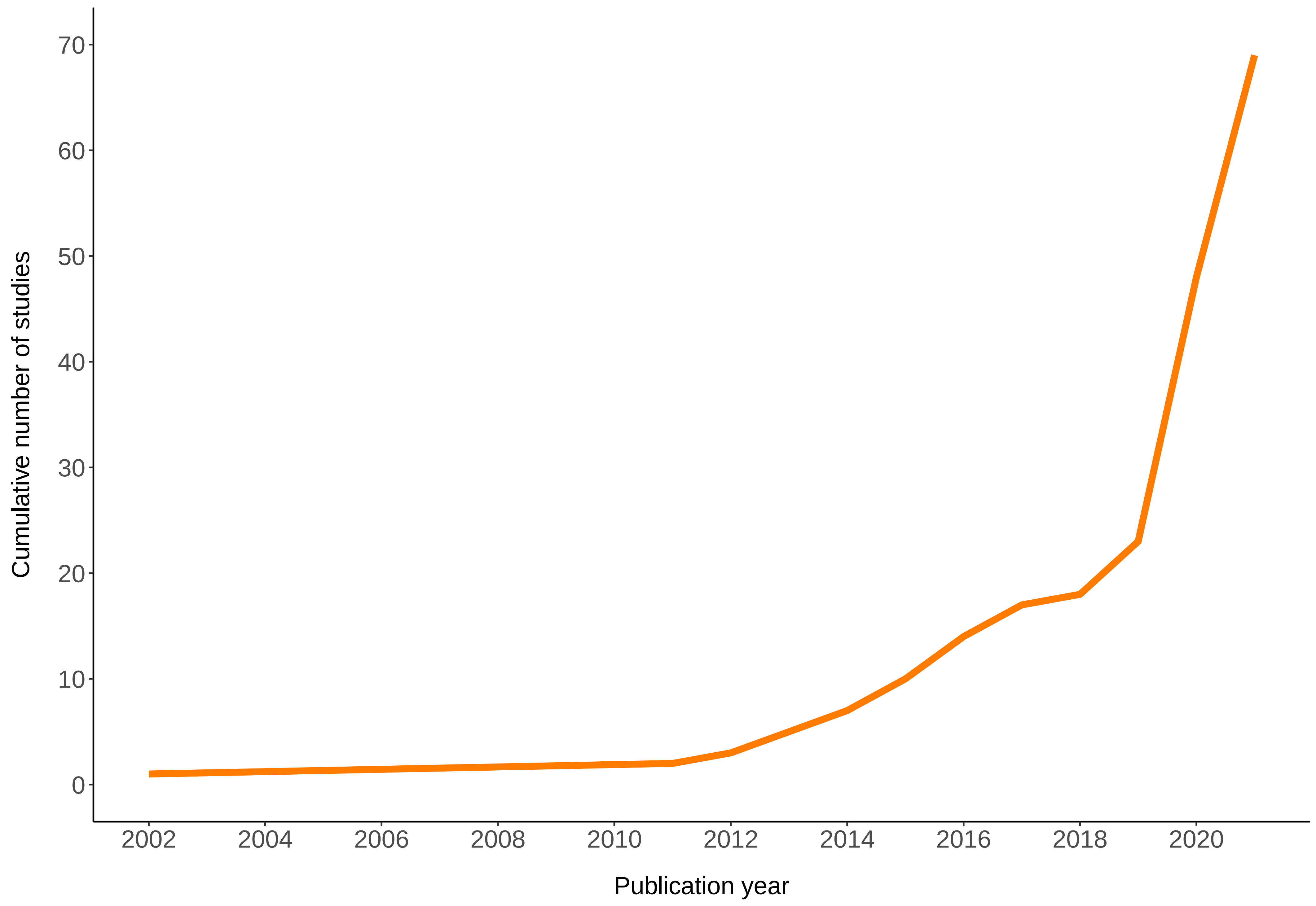

**Supplementary Figure1: Cumulative number of epigenome-wide studies using ML methods to identify DNA methylation biomarkers associated with cancer prognosis per year**

**Supplementary Table1: PRISMA-ScR Checklist**

| SECTION | ITEM | PRISMA-ScR CHECKLIST ITEM | REPORTED ON PAGE # |
|---|---|---|---|
| **TITLE** | | | |
| Title | 1 | Identify the report as a scoping review. | p1 |
| **ABSTRACT** | | | |
| Structured summary | 2 | Provide a structured summary that includes (as applicable): background, objectives, eligibility criteria, sources of evidence, charting methods, results, and conclusions that relate to the review questions and objectives. | p2 |
| **INTRODUCTION** | | | |
| Rationale | 3 | Describe the rationale for the review in the context of what is already known. Explain why the review questions/objectives lend themselves to a scoping review approach. | p4-5 |
| Objectives | 4 | Provide an explicit statement of the questions and objectives being addressed with reference to their key elements (e.g., population or participants, concepts, and context) or other relevant key elements used to conceptualize the review questions and/or objectives. | p5 |
| **METHODS** | | | |
| Protocol and registration | 5 | Indicate whether a review protocol exists; state if and where it can be accessed (e.g., a Web address); and if available, provide registration information, including the registration number. | p6 |
| Eligibility criteria | 6 | Specify characteristics of the sources of evidence used as eligibility criteria (e.g., years considered, language, and publication status), and provide a rationale. | p6-7 |
| Information sources | 7 | Describe all information sources in the search (e.g., databases with dates of coverage and contact with authors to identify additional sources), as well as the date the most recent search was executed. | p6 |
| Search | 8 | Present the full electronic search strategy for at least 1 database, including any limits used, such that it could be repeated. | p6 |
| Selection of sources of evidence | 9 | State the process for selecting sources of evidence (i.e., screening and eligibility) included in the scoping review. | p6 |
| Data charting process | 10 | Describe the methods of charting data from the included sources of evidence (e.g., calibrated forms or forms that have been tested by the team before their use, and whether data charting was done independently or in duplicate) and any processes for obtaining and confirming data from investigators. | p8 |
| Data items | 11 | List and define all variables for which data were sought and any assumptions and simplifications made. | p8 |

| | | | |
|---|---|---|---|
| Critical appraisal of individual sources of evidence | 12 | If done, provide a rationale for conducting a critical appraisal of included sources of evidence; describe the methods used and how this information was used in any data synthesis (if appropriate). | p7-8 |
| Synthesis of results | 13 | Describe the methods of handling and summarizing the data that were charted. | p8 |
| **RESULTS** | | | |
| Selection of sources of evidence | 14 | Give numbers of sources of evidence screened, assessed for eligibility, and included in the review, with reasons for exclusions at each stage, ideally using a flow diagram. | Figure1 |
| Characteristics of sources of evidence | 15 | For each source of evidence, present characteristics for which data were charted and provide the citations. | Table1, p9-10 |
| Critical appraisal within sources of evidence | 16 | If done, present data on critical appraisal of included sources of evidence (see item 12). | Figure2, supplementary table3, p10 |
| Results of individual sources of evidence | 17 | For each included source of evidence, present the relevant data that were charted that relate to the review questions and objectives. | Supplementary table 4-6 |
| Synthesis of results | 18 | Summarize and/or present the charting results as they relate to the review questions and objectives. | p11-15 |
| **DISCUSSION** | | | |
| Summary of evidence | 19 | Summarize the main results (including an overview of concepts, themes, and types of evidence available), link to the review questions and objectives, and consider the relevance to key groups. | p15 |
| Limitations | 20 | Discuss the limitations of the scoping review process. | p19 |
| Conclusions | 21 | Provide a general interpretation of the results with respect to the review questions and objectives, as well as potential implications and/or next steps. | p20 |
| **FUNDING** | | | |
| Funding | 22 | Describe sources of funding for the included sources of evidence, as well as sources of funding for the scoping review. Describe the role of the funders of the scoping review. | p21 |

**Supplementary Table 2: Search strategy**

| Keywords | | Pubmed | EMBASE | Web of science |
|---|---|---|---|---|
| AI | #1 | ("machine learning"[MeSH Terms] OR "artificial intelligence"[MeSH Terms] OR "decision support"[Text Word] OR "clustering"[Text Word] OR "deep learning"[Text Word] OR "neural network*"[Text Word]) AND ((journalarticle[Filter]) AND (humans[Filter]) | ('machine learning'/exp OR 'machine learning' OR 'artificial intelligence'/exp OR 'artificial intelligence' OR 'decision support'/exp OR 'decision support' OR 'clustering' OR 'deep learning'/exp OR 'deep learning' OR 'neural network*') AND [article]/lim AND [humans]/lim AND [english]/lim AND [embase]/lim AND [1990-2021]/py | (((((ALL= (machine learning)) OR ALL=(artificial intelligence)) OR ALL=(decision support)) OR ALL=(clustering)) OR ALL=(deep learning)) OR ALL=(neural network*) |
| Cancer | #2 | ("neoplasms"[MeSH Terms] OR "cancer"[Title/Abstract] OR "tumor*"[Title/Abstract] OR "carcinoma*"[Title/Abstract] OR "pan-cancer"[Title/Abstract] OR "adenocarcinoma*"[Title/Abstract] OR "leukemia"[Title/Abstract]) | (('neoplasm':ab,ti OR 'cancer':ab,ti OR 'tumor*':ab,ti) AND 'carcinoma':ab,ti OR 'pan cancer':ab,ti OR 'adenocarcinoma*':ti,ab OR 'leukemia':ab,ti) AND [article]/lim AND [humans]/lim AND [english]/lim AND [embase]/lim AND [1990-2021]/py | ((((((TI=(neoplasms)) OR TI=(cancer)) OR TI=(tumor)) OR TI=(carcinoma)) OR TI=(pan-cancer)) OR TI=(adenocarcinoma)) OR TI=(leukemia) |
| Epigenetic signatures | #3 | ("methylation"[MeSH Terms] OR "dna methylation*"[MeSH Terms] OR "methylat*"[Title/Abstract] OR "epigenetic*"[Title/Abstract] OR "epigenetic marker*"[Title/Abstract] OR "DNA-methylation"[Title/Abstract] OR "hypermethylat*"[Title/Abstract] OR "hypomethylat*"[Title/Abstract] OR "CpG"[Title/Abstract] OR "epigenome-wide"[Title/Abstract] OR "methylation-based"[Title/Abstract]))) | ('methylation':ab OR 'dna methylation':ab OR 'methylat*':ab OR 'epigenetic*':ab OR 'epigenetic marker*':ab OR 'dna-methylation':ab OR 'hypermethylat*':ab OR 'hypomethylat*':ab OR 'cpg':ab OR 'epigenome-wide':ab OR 'methylation-based':ab) AND [article]/lim AND [humans]/lim AND [english]/lim AND [embase]/lim AND [1990-2021]/py | ((((((((((AB=(methylation)) OR AB=(dna methylation)) OR AB=(methylat*)) OR AB=(epigenetic*)) OR AB=(epigenetic marker*)) OR AB=(DNA-methylation)) OR AB=(hypermethylat*)) OR AB=(hypomethylat*)) OR AB=(CpG)) OR AB=(epigenome-wide)) OR AB=(methylation-based) |
| Prognosis | #4 | ("mortality"[MeSH Terms] OR "survival"[MeSH Terms] OR "survival analysis"[MeSH Terms] OR "mortality"[Title/Abstract] OR "survival"[Title/Abstract] OR "prognos*"[Title/Abstract] OR "predict*"[Title/Abstract] OR "outcome*"[Title/Abstract] OR "recurr*"[Title/Abstract]) | (('mortality':ab,ti OR 'survival':ti,ab OR 'survival analysis':ti,ab) AND 'prognos*':ti,ab OR 'predict*':ti,ab OR 'outcome*':ti,ab OR 'recurr*':ti,ab) AND [article]/lim AND [humans]/lim AND [english]/lim AND [embase]/lim AND [1990-2021]/py | ((((((TI=(mortality)) OR TI=(survival)) OR TI=(survival analysis)) OR TI=(prognos*)) OR TI=(predict*)) OR TI=(outcome*)) OR TI=(recurr*) |
| Final | #5 | #1 AND #2 AND #3 AND #4 AND ((journalarticle[Filter]) AND (humans[Filter]) AND (1990/1/1:2021/9[pdat]) AND (english[Filter])) | #1 AND #2 AND #3 AND #4 | (#1 AND #2 AND #3 AND #4) AND Articles (Documents Types) AND English (Languages) AND DOP = (1990-01-01 -2021) |

**Supplementary Table 3: Characteristics of included studies**

| Author, year | Study characteristics | | | | | | | | Methodological characteristics | | | | | | | | Performance measurement | | | |
|---|---|---|---|---|---|---|---|---|---|---|---|---|---|---|---|---|---|---|---|---|
| | | | | | | | | | | | | | | | | | Discrimination | | | |
| | Geographic origin | Median follow up (yr) | Cancer type | Cancer stage | Outcome | Type of biospecimen | Type of methylation array | Type of methylation signatures | Size of training set | Source of training set | Siz of internal validation | Size of external validation | Source of external validation data | Handling of missing values | ML methods used | PI construction | Cox regression | C-index | ROC/AUC | Calibration |
| Wei et al,[1] 2002 | Multi | NR | Ovary | III-IV | PFS | Tumor tissue | Differential methylation hybridization | CpGs | 19 | Clinical cohort | NP | NP | NP | NR | Hierarchical clustering Batch sequential forward selection | ✗ | ✓ | ✗ | ✗ | ✗ |
| Walker et al,[2] 2011 | UK | NR | Multiple myeloma | I-III | OS | Bone marrow | Illumina HM 27K | CpGs | 161 | Clinical cohort | NP | NP | NP | Row means | Hierarchical clustering | ✗ | ✗ | ✗ | ✗ | ✗ |
| Sigalotti et al,[3] 2012 | Italy | NR | Cutaneous melanoma | III | OS | Tumor tissue | Illumina HM 27K | Genes | 45 | Clinical cohort | NP | NP | NP | NR | K-means clustering Nearest shrunken centroid algorithm | ✗ | ✗ | ✗ | ✗ | ✗ |
| Chambwe et al,[4] 2014 | Canada | NR | Non-Hodgkin lymphoma | I-IV | OS | Tumor tissue | HELP Assay | Genes | 140 | Clinical cohort | NP | NP | NP | NR | Hierarchical clustering Consensus clustering | ✗ | ✗ | ✗ | ✗ | ✗ |
| Fleischer et al,[5] 2014 | Norway | NR | Breast | I-IV | Cancer-specific survival | Tumor tissue | Illumina HM 450K | CpGs, Genes | 176 | Clinical cohort | NP | 583 | TCGA | NR | LASSO | ✓ | ✓ | ✗ | ✗ | ✗ |
| Mah et al,[6] 2014 | Singapore | NR | Liver | I-III | OS and RFS | Tumor tissue | Illumina HM 27K | CpGs, Genes | 59 | Clinical cohort | NP | NP | NP | NR | Consensus clustering | ✗ | ✓ | ✗ | ✗ | ✗ |
| Wang et al,[7] 2014 | US | NR | Ovary | I-IV | RFS | Tumor tissue | Illumina HM 450K | CpGs | 168 | Clinical cohort | 169 | NP | NP | NR | Recursive partitioning mixture model Bayesian network classifier | ✗ | ✓ | ✗ | ✗ | ✗ |
| Ouchi et al,[8] 2015 | Japan | NR | Colon and rectum | I-IV | Response to therapy | Tumor tissue | Illumina HM 450K | CpGs | 45 | Clinical cohort | 52 | NP | NP | NR | Hierarchical clustering Random forest | ✗ | ✗ | ✗ | ✗ | ✗ |
| Villanueva et al,[9] 2015 | Multi | 4 | Liver | NR | OS and RFS | Tumor tissue | Illumina HM 450K | CpGs | 221 | Clinical cohort | NP | 83 | Clinical cohort | NR | Random survival forests | ✓ | ✓ | ✗ | ✗ | ✗ |
| Wei et al,[10] 2015 | China | NR | Kidney | I-IV | OS | Tumor tissue | Illumina HM 450K | CpGs, Genes | 168 | Clinical cohort | NP | 243, 284, 298 | TCGA and clinical cohorts | NR | LASSO | ✓ | ✓ | ✗ | ✗ | ✗ |
| Bjaanaes et al,[11] 2016 | Norway | 3.7 | Lung | I-IV | PFS | Tumor tissue | Illumina HM 450K | CpGs | 164 | Clinical cohort | NP | 244 | TCGA | NR | LASSO | ✓ | ✓ | ✗ | ✗ | ✗ |
| Lim et al,[12] 2016 | Multi | NR | Oral tongue | I-IV | OS | Tumor tissue | Illumina HM 450K | CpGs | 174 | Clinical cohort, TCGA | NP | NP | NP | NR | Centroid feature selection SVM-RFE | ✓ | ✗ | ✓ | ✗ | ✗ |

| | | | | | | | | | | | | | | | | | | | | |
|---|---|---|---|---|---|---|---|---|---|---|---|---|---|---|---|---|---|---|---|---|
| Lu et al,[13] 2016 | Multi | NR | Glioblastoma | NR | Long-term survival | Tumor tissue | Illumina HM 450K and 27K | CpGs | 283 | TCGA | NP | NP | NP | Median | LASSO | ✓ | ✕ | ✕ | ✓ | ✕ |

| **Author, year** | **Study characteristics** | | | | | | | | **Methodological characteristics** | | | | | | | | **Performance measurement** | | | |
|---|---|---|---|---|---|---|---|---|---|---|---|---|---|---|---|---|---|---|---|---|
| | | | | | | | | | | | | | | | | | **Discrimination** | | | **Calibration** |
| | **Geographic origin** | **Median follow up (yr)** | **Cancer type** | **Cancer stage** | **Outcome** | **Type of biospecimen** | **Type of methylation array** | **Type of methylation signatures** | **Size of training set** | **Source of training set** | **Siz of internal validation** | **Size of external validation** | **Source of external validation data** | **Handling of missing values** | **ML methods used** | **PI construction** | **Cox regression** | **C-index** | **ROC/AUC** | |
| Saito et al,[14] 2016 | Japan | 3.1 | Lung | I-IV | RFS | Tumor tissue | Illumina HM 27K | CpGs, Genes | 28 | Clinical cohort | NP | NP | NP | NR | Hierarchical clustering Consensus clustering | ✕ | ✕ | ✕ | ✕ | ✕ |
| Hao et al,[15] 2017 | Multi | NR | Lung, Breast | I-IV | OS | Tumor tissue | Illumina HM 450K | CpGs | Lung: 585 Breast: 520 | TCGA | Lung: 254 Breast: 270 | NP | NP | NR | LASSO Boosting | ✓ | ✕ | ✓ | ✕ | ✕ |
| Qiu et al,[16] 2017 | China | 4.6 | Liver | I | OS and RFS | Tumor tissue | Illumina HM 450K | CpGs | 206 | Clinical cohort | 141 | 191, 104, 125 | 2 clinical cohorts and TCGA | K-nearest neighor | LASSO SVM-RFE | ✓ | ✓ | ✓ | ✓ | ✓ |
| Stieglitz et al,[17] 2017 | Multi | NR | Leukemia | NR | PFS | Bone marrow, splenic tissue or peripheral blood | Illumina HM 450K | CpGs | 39 | Clinical cohort | NP | NP | NP | NR | Hierarchical clustering | ✕ | ✕ | ✕ | ✕ | ✕ |
| Zhang et al,[18] 2018 | Multi | NR | Breast | I-IV | OS | Tumor tissue | Illumina HM 450K | CpGs | 335 | TCGA | NP | 119 | GEO | K-nearest neighor | Consensus clustering Bayesian network classifier | ✕ | ✕ | ✕ | ✕ | ✕ |
| Chen et al,[19] 2019 | Multi | NR | Kidney | I-IV | OS | Tumor tissue | Illumina HM 450K | CpGs | 153 | TCGA | 154 | NP | NP | K-nearest neighor | Consensus clustering Bayesian network classifier | ✕ | ✕ | ✕ | ✕ | ✕ |
| Dong et al,[20] 2019 | Multi | NR | Liver | NR | OS | Tumor tissue | Illumina HM 450K | CpGs | 130 | TCGA | 33 | 19 | GEO | NR | SVM-RFE FW-SVM | ✓ | ✓ | ✕ | ✕ | ✕ |
| Li et al,[21] 2019 | Multi | NR | Cutaneous melanoma | I-IV | OS | Tumor tissue | Illumina HM 450K | CpGs, Genes | 447 | TCGA | NP | NP | NP | NR | Consensus clustering | ✕ | ✕ | ✕ | ✕ | ✕ |
| Wang et al,[22] 2019 | Multi | NR | Lung | NR | OS | Tumor tissue | Illumina HM 450K | CpGs | 446 | TCGA | NP | NP | NP | NR | LASSO RFE | ✓ | ✓ | ✕ | ✓ | ✓ |
| Yang et al,[23] 2019 | Multi | NR | Colon | I-IV | OS | Tumor tissue | Illumina HM 450K and 27K | CpGs | 272 | TCGA | 187 | NP | NP | K-nearest neighor | Consensus clustering | ✓ | ✓ | ✕ | ✕ | ✕ |
| Bian et al,[24] 2020 | Multi | NR | Stomach | I-IV | OS | Tumor tissue | Illumina HM 450K | CpGs | 183 | TCGA | 183 | NP | NP | K-nearest neighor | Consensus clustering LASSO | ✓ | ✓ | ✕ | ✓ | ✕ |
| Cai et al,[25] 2020 | Multi | NR | Lung | I-IV | OS | Tumor tissue | Illumina HM 450K and 27K | CpGs | 405 | TCGA | 62 | NP | NP | K-nearest neighor | Consensus clustering | ✓ | ✓ | ✓ | ✓ | ✓ |

| | | | | | | | | | | | | | | | | | | | | |
|---|---|---|---|---|---|---|---|---|---|---|---|---|---|---|---|---|---|---|---|---|
| Chen et al,[26] 2020 | Multi | NR | Thyroid | I-IV | RFS | Tumor tissue | Illumina HM 450K | CpGs | 383 | TCGA | 163 | NP | NP | K-nearest neighor | LASSO | ✓ | ✓ | ✓ | ✓ | ✓ |

| Author, year | Study characteristics | | | | | | | | Methodological characteristics | | | | | | | | Performance measurement | | | |
|---|---|---|---|---|---|---|---|---|---|---|---|---|---|---|---|---|---|---|---|---|
| | | | | | | | | | | | | | | | | | Discrimination | | | Calibration |
| | Geographic origin | Median follow up (yr) | Cancer type | Cancer stage | Outcome | Type of biospecimen | Type of methylation array | Type of methylation signatures | Size of training set | Source of training set | Siz of internal validation | Size of external validation | Source of external validation data | Handling of missing values | ML methods used | PI construction | Cox regression | C-index | ROC/AUC | |
| Deng et al,[27] 2020 | Multi | NR | Colon and rectum | I-IV | PFS | Tumor tissue | Illumina HM 450K | CpGs | 189 | TCGA | 188 | NP | NP | NR | LASSO<br>SVM-RFE | ✓ | ✓ | ✓ | ✓ | ✓ |
| Dong et al,[28] 2020 | Multi | NR | Lung | I-IV | OS | Tumor tissue | Illumina HM 450K | CpGs | 267 | TCGA | 134 | 97 | GEO | NR | Random survival forests-variable hunting | ✓ | ✓ | ✗ | ✓ | ✓ |
| He et al,[29] 2020 | Multi | NR | Liver | NR | Survival | Tumor tissue | Illumina HM 450K | CpGs | 174 | TCGA | 174 | NP | NP | K-nearest neighor | Consensus clustering<br>Bayesian network classifier | ✗ | ✓ | ✗ | ✗ | ✗ |
| Lee et al,[30] 2020 | Multi | NR | Lung | I-III | OS | Tumor tissue | Illumina HM 450K | Genes | 399 | TCGA | NP | 198, 32, 35, 94 | GEO, TCGA | Exclusion | Autoencoder<br>LASSO<br>K-means clustering<br>Random forest | ✗ | ✗ | ✗ | ✗ | ✗ |
| Li, C et al,[31] 2020 | Multi | NR | Cervix | NR | OS | Tumor tissue | Illumina HM 450K | CpGs | 137 | TCGA | 138 | NP | NP | K-nearest neighor | Consensus clustering<br>LASSO | ✓ | ✓ | ✗ | ✗ | ✗ |
| Li, T et al,[32] 2020 | Multi | NR | Stomach | I-IV | OS | Tumor tissue | Illumina HM 450K and 27K | CpGs | 235 | TCGA | 235 | NP | NP | Exclusion | Consensus clustering | ✗ | ✓ | ✗ | ✗ | ✗ |
| Lian et al,[33] 2020 | Multi | NR | Stomach | NR | OS | Tumor tissue | Illumina HM 450K | CpGs | 197 | TCGA | 197 | NP | NP | K-nearest neighor | Consensus clustering<br>Random forest | ✗ | ✓ | ✗ | ✗ | ✗ |
| Luo, H et al,[34] 2020 | Multi | 2.2 | Colon and rectum | I-IV | OS | Blood | Illumina HM 450K | CpGs | 528 | Clinical cohort, TCGA | 273 | NP | NP | Exclusion | LASSO | ✓ | ✓ | ✓ | ✓ | ✓ |
| Luo, R et al,[35] 2020 | Multi | NR | Lung | I-IV | RFS | Tumor tissue | Illumina HM 450K | CpGs | 823 | TCGA | NP | 444, 164, 60 | GEO | NR | LASSO<br>Random forest | ✓ | ✓ | ✗ | ✓ | ✗ |
| Lv et al,[36] 2020 | Multi | NR | Colon | I-IV | OS | Tumor tissue | Illumina HM 450K | Genes | 288 | TCGA | NP | NP | NP | Imputation | Autoencoder<br>K-means clustering<br>Support vector machine | ✗ | ✗ | ✗ | ✗ | ✗ |
| Ma, H et al,[37] 2020 | Multi | NR | Glioblastoma | NR | OS and RFS | Tumor tissue | Illumina HM 450K | CpGs | 69 | TCGA | 69 | 12 | Chinese Glioma Genome Altas database | K-nearest neighor | Consensus clustering | ✗ | ✓ | ✗ | ✗ | ✗ |
| Ma, X et al,[38] 2020 | Multi | NR | Stomach | I-IV | OS | Tumor tissue | Illumina HM 450K | CpGs | 268 | TCGA | 114 | NP | NP | K-nearest neighor | LASSO | ✓ | ✓ | ✓ | ✓ | ✓ |

| Author, year | Study characteristics | | | | | | | | Methodological characteristics | | | | | | | | Performance measurement | | | |
|---|---|---|---|---|---|---|---|---|---|---|---|---|---|---|---|---|---|---|---|---|
| | | | | | | | | | | | | | | | | | Discrimination | | | |
| | Geographic origin | Median follow up (yr) | Cancer type | Cancer stage | Outcome | Type of biospecimen | Type of methylation array | Type of methylation signatures | Size of training set | Source of training set | Siz of internal validation | Size of external validation | Source of external validation data | Handling of missing values | ML methods used | PI construction | Cox regression | C-index | ROC/AUC | Calibration |
| Macías-García et al,[39] 2020 | Multi | NR | Breast | NR | RFS | Tumor tissue | Illumina HM 450K | Genes | 99 | TCGA | NP | NP | NP | Exclusion | Autoencoder | ✕ | ✕ | ✕ | ✕ | ✕ |
| Shi et al,[40] 2020 | Multi | NR | Lung | I-IV | OS | Tumor tissue | Illumina HM 450K and 27K | CpGs | 335 | TCGA | 150 | NP | NP | K-nearest neighor | Consensus clustering | ✕ | ✓ | ✕ | ✓ | ✕ |
| Tian et al,[41] 2020 | Multi | NR | Bladder | I-IV | OS | Tumor tissue | NR | CpGs | 203 | TCGA | 204 | NP | NP | K-nearest neighor | Consensus clustering | ✓ | ✓ | ✓ | ✓ | ✓ |
| Wang, X et al,[42] 2020 | Multi | NR | Colon and rectum | I-IV | OS, PFS | Tumor tissue | Illumina HM 450K | CpGs | 249 | TCGA | 129 | NP | NP | NR | Hierarchical clustering LASSO | ✓ | ✓ | ✓ | ✓ | ✓ |
| Wang, Y et al,[43] 2020 | Multi | NR | Lung | I-IV | OS | Tumor tissue | Illumina HM 450K | CpGs | 174 | TCGA | 175 | NP | NP | K-nearest neighor | Consensus clustering | ✕ | ✓ | ✕ | ✕ | ✕ |
| Xiang et al,[44] 2020 | Multi | NR | Stomach, Colon | I-IV | OS | Tumor tissue | Illumina HM 450K and 27K | CpGs | Stomach : 406 Colon: 385 | TCGA | NP | NR | the UCSC Xena platform | NR | Hierarchical clustering LASSO | ✓ | ✓ | ✓ | ✓ | ✓ |
| Yu et al,[45] 2020 | Multi | NR | Esophagus | I-IV | OS | Tumor tissue | Illumina HM 450K | Genes | 96 | TCGA | NP | NP | NP | Imputation | Autoencoder K-means clustering Support vector machine | ✕ | ✕ | ✕ | ✕ | ✕ |
| Zhang, C et al,[46] 2020 | Multi | NR | Kidney | I-IV | OS | Tumor tissue | Illumina HM 450K | Genes | 306 | TCGA | NP | 102 | European Bioinformatics Institute | NR | LASSO | ✓ | ✓ | ✕ | ✓ | ✓ |
| Zhang, E et al,[47] 2020 | Multi | NR | Prostate | I-IV | OS | Tumor tissue | Illumina HM 450K | CpGs, Genes | 201 | TCGA | 100 | NP | NP | NR | LASSO | ✕ | ✓ | ✕ | ✓ | ✕ |
| Zheng et al,[48] 2020 | Multi | NR | Leukemia | NR | PFS | Tumor tissue | Illumina HM 27K | CpGs, Genes | 219 | TCGA | NP | NP | NP | NR | Consensus clustering | ✕ | ✕ | ✕ | ✓ | ✕ |
| Bedon et al,[49] 2021 | Multi | NR | Liver | NR | PFS | Tumor tissue | Illumina HM 450K | CpGs | 300 | TCGA | 74 | NP | NP | NR | Recursive Features Selection All relevant features analysis (Boruta) Random forest | ✕ | ✓ | ✕ | ✕ | ✕ |
| Chen et al,[50] 2021 | Multi | NR | Esophagus | NR | Survival | Tumor tissue | Illumina HM 450K | CpGs | 89 | TCGA | 89 | NP | NP | K-nearest neighor | Consensus clustering | ✕ | ✓ | ✕ | ✕ | ✕ |
| Hao et al,[51] 2021 | Multi | NR | Liver | I-IV | OS | Tumor tissue | Illumina HM 450K | Genes | 296 | GEO, TCGA | 148 | NP | NP | K-nearest neighor | LASSO | ✕ | ✓ | ✕ | ✓ | ✕ |

| Author, year | Study characteristics | | | | | | | | Methodological characteristics | | | | | | | | Performance measurement | | | |
|---|---|---|---|---|---|---|---|---|---|---|---|---|---|---|---|---|---|---|---|---|
| | | | | | | | | | | | | | | | | | Discrimination | | | |
| | Geographic origin | Median follow up (yr) | Cancer type | Cancer stage | Outcome | Type of biospecimen | Type of methylation array | Type of methylation signatures | Size of training set | Source of training set | Siz of internal validation | Size of external validation | Source of external validation data | Handling of missing values | ML methods used | PI constructio n | Cox regression | C-index | ROC/AUC | Calibration |
| Huang, G et al,[52] 2021 | Multi | NR | Lung | I-IV | OS | Tumor tissue | Illumina HM 450K | CpGs | 274 | TCGA | NP | NP | NP | K-nearest neighor | Consensus clustering | ✗ | ✓ | ✗ | ✓ | ✗ |
| Huang, GJ et al,[53] 2021 | Multi | NR | Liver | I-IV | OS and RFS | Tumor tissue | Illumina HM 450K | Genes | 364 | TCGA | NP | 82, 41, 41 | GEO | imputation | Bidirectional deep neural networks<br>K-means clustering<br>Support vector machine | ✗ | ✗ | ✗ | ✗ | ✗ |
| Huang, H et al,[54] 2021 | Multi | NR | Colon and rectum | I-IV | OS | Tumor tissue | Illumina HM 450K | Genes | 367 | TCGA | NP | 355 | GEO | NR | LASSO | ✓ | ✓ | ✓ | ✓ | ✓ |
| Huang, W et al,[55] 2021 | Multi | NR | Pancreas | I-IV | OS | Tumor tissue | Illumina HM 450K | Genes | 185 | TCGA | NP | 273, 65 | Clinical cohort and GEO | NR | LASSO | ✓ | ✓ | ✓ | ✓ | ✓ |
| Lee et al,[56] 2021 | Multi | NR | Lung | I-IV | OS | Tumor tissue | Illumina HM 450K | CpGs | 187 | GEO, TCGA | 92 | 23 | GEO | NR | Random survival forests-variable hunting | ✓ | ✓ | ✗ | ✓ | ✗ |
| Li, K et al,[57] 2021 | Multi | NR | Soft tissue sarcoma | NR | OS | Tumor tissue | Illumina HM 450K | CpGs | 269 | TCGA | NP | NP | NP | K-nearest neighor | Consensus clustering<br>LASSO | ✓ | ✓ | ✗ | ✓ | ✗ |
| Li, X et al,[58] 2021 | Multi | NR | Lung | I-IV | OS | Tumor tissue | Illumina HM 450K and 27K | CpGs | 378 | TCGA | 126 | NP | NP | NR | Consensus clustering | ✓ | ✓ | ✗ | ✓ | ✗ |
| Peng et al,[59] 2021 | Multi | NR | Colon | I-IV | OS | Tumor tissue | Illumina HM 450K | Genes | 141 | TCGA | 140 | NP | NP | NR | LASSO | ✓ | ✓ | ✗ | ✓ | ✗ |
| Shang et al,[60] 2021 | Multi | NR | Lung | NR | Response to therapy | Tumor tissue | NR | CpGs | 78 | GEO | NP | 486, 370 | TCGA | NR | LASSO<br>Random forest<br>Support vector machine | ✓ | ✗ | ✗ | ✓ | ✗ |
| Sun et al,[61] 2021 | Multi | NR | Colon | I-IV | OS | Tumor tissue | Illumina HM 450K | Genes | 142 | TCGA | 143 | NP | NP | NR | LASSO | ✓ | ✓ | ✓ | ✓ | ✗ |
| Tan et al,[62] 2021 | Multi | NR | Colon and rectum | NR | OS | Tumor tissue | Illumina HM 27K | Genes | 556 | GEO | NP | 469 | TCGA | NR | LASSO | ✓ | ✓ | ✗ | ✗ | ✗ |
| Wang, L et al,[63] 2021 | Multi | NR | Endometri um | I-IV | Survival | Tumor tissue | Illumina HM 450K and 27K | CpGs | 482 | TCGA | 118 | NP | NP | K-nearest neighor | Consensus clustering | ✓ | ✓ | ✗ | ✓ | ✗ |
| Wang, T et al,[64] 2021 | Multi | NR | Prostate | NR | RFS | Tumor tissue | Illumina HM 450K | Genes | 494 | TCGA | NP | NP | NP | Exclusion | Autoencoder<br>K-means clustering | ✓ | ✗ | ✓ | ✓ | ✗ |
| Wu et al,[65] 2021 | Multi | NR | Breast | I-IV | OS | Tumor tissue | Illumina HM 450K and 27K | CpGs | 890 | TCGA | 342 | NP | NP | K-nearest neighor | Consensus clustering | ✗ | ✓ | ✗ | ✓ | ✗ |

| Author, year | Study characteristics | | | | | | | | Methodological characteristics | | | | | | | | Performance measurement | | | |
|---|---|---|---|---|---|---|---|---|---|---|---|---|---|---|---|---|---|---|---|---|
| | | | | | | | | | | | | | | | | | Discrimination | | | Calibration |
| | Geographic origin | Median follow up (yr) | Cancer type | Cancer stage | Outcome | Type of biospecimen | Type of methylation array | Type of methylation signatures | Size of training set | Source of training set | Siz of internal validation | Size of external validation | Source of external validation data | Handling of missing values | ML methods used | PI constructio n | Cox regression | C-index | ROC/AUC | |
| Xu et al,[66] 2021 | Multi | NR | Liver | I-IV | OS | Tumor tissue | Illumina HM 450K | Genes | 114 | TCGA | 112 | 260 | ICGC | NR | Consensus clustering<br>LASSO | ✓ | ✓ | × | ✓ | ✓ |
| Yin, L et al,[67] 2021 | Multi | NR | Ovary | I-IV | OS | Tumor tissue | Illumina HM 27K | CpGs | 285 | TCGA | 286 | NP | NP | K-nearest neighor | Consensus clustering | × | ✓ | × | × | × |
| Yin, X et al,[68] 2021 | Multi | NR | Pancreas | I-IV | OS | Tumor tissue | Illumina HM 450K | CpGs | 125 | TCGA | 53 | NP | NP | K-nearest neighor | Consensus clustering<br>LASSO | ✓ | ✓ | ✓ | ✓ | ✓ |
| Yu et al,[69] 2021 | Multi | NR | Lung | I-IV | RFS | Tumor tissue | Illumina HM 450K | CpGs | 455 | TCGA | NP | 155 | GEO | Row means | LASSO<br>Elastic-net<br>Support vector machine<br>Stacking ensemble model | ✓ | ✓ | × | ✓ | × |

**Supplementary Table 4: Quality assessment for each included study**

| Study | Was the follow up > 3 years? (Yes=1, No/NR=0) | Was the sample size of training dataset at least 100? (Yes=1, No/NR=0) | Were there appropriate methods of imputation used for missing data? (Yes=1, No/NR=0) | Were methylation biomarkers selected or assessed by adjusting for age, sex, and tumor stage? (Yes=1, No/NR=0) | Was validation performed? (No =0, Internal =1, External =2) | Were relevant performance measures evaluated appropriately (No=0, only discrimination=1, both discrimination and calibration=2) | Was Cross-validation or Bootstrapping used to evaluate performance (Yes =1, No/NR=0) | Total score | Quality |
|---|---|---|---|---|---|---|---|---|---|
| Wei et al,[1] 2002 | 1 | 0 | 0 | 0 | 0 | 1 | 0 | 2 | Low |
| Walker et al,[2] 2011 | 1 | 1 | 1 | 0 | 0 | 1 | 0 | 4 | Medium |
| Sigalotti et al,[3] 2012 | 1 | 0 | 0 | 1 | 0 | 1 | 0 | 3 | Low |
| Chambwe et al,[4] 2014 | 1 | 1 | 0 | 0 | 0 | 1 | 0 | 3 | Low |
| Fleischer et al,[5] 2014 | 1 | 1 | 0 | 0 | 2 | 1 | 0 | 5 | Medium |
| Mah et al,[6] 2014 | 1 | 0 | 0 | 0 | 0 | 1 | 0 | 2 | Low |
| Wang et al,[7] 2014 | 1 | 1 | 0 | 1 | 1 | 1 | 0 | 5 | Medium |
| Ouchi et al,[8] 2015 | 0 | 0 | 0 | 0 | 1 | 0 | 0 | 1 | Low |
| Villanueva et al,[9] 2015 | 1 | 1 | 0 | 0 | 2 | 1 | 1 | 6 | Medium |
| Wei et al,[10] 2015 | 1 | 1 | 0 | 1 | 2 | 1 | 0 | 6 | Medium |
| Bjaanaes et al,[11] 2016 | 1 | 1 | 0 | 1 | 2 | 1 | 0 | 6 | Medium |
| Lim et al,[12] 2016 | 0 | 1 | 0 | 0 | 0 | 0 | 1 | 2 | Low |
| Lu et al,[13] 2016 | 1 | 1 | 1 | 0 | 1 | 1 | 1 | 6 | Medium |
| Saito et al,[14] 2016 | 1 | 0 | 0 | 1 | 0 | 1 | 0 | 3 | Low |
| Hao et al,[15] 2017 | 1 | 1 | 0 | 0 | 1 | 1 | 0 | 4 | Medium |
| Qiu et al,[16] 2017 | 1 | 1 | 1 | 0 | 2 | 2 | 0 | 7 | High |
| Stieglitz et al,[17] 2017 | 1 | 0 | 0 | 0 | 2 | 1 | 0 | 4 | Medium |
| Zhang et al,[18] 2018 | 1 | 1 | 1 | 1 | 2 | 1 | 1 | 8 | High |
| Chen et al,[19] 2019 | 1 | 1 | 1 | 0 | 1 | 1 | 1 | 6 | Medium |
| Dong et al,[20] 2019 | 1 | 1 | 0 | 0 | 2 | 1 | 1 | 6 | Medium |
| Li et al,[21] 2019 | 1 | 1 | 0 | 0 | 0 | 1 | 0 | 3 | Low |
| Wang et al,[22] 2019 | 1 | 1 | 0 | 1 | 1 | 2 | 1 | 7 | High |
| Yang et al,[23] 2019 | 1 | 1 | 1 | 0 | 1 | 1 | 0 | 5 | Medium |
| Bian et al,[24] 2020 | 1 | 1 | 1 | 1 | 1 | 1 | 0 | 6 | Medium |
| Cai et al,[25] 2020 | 1 | 1 | 1 | 1 | 1 | 2 | 0 | 7 | High |
| Chen et al,[26] 2020 | 1 | 1 | 1 | 0 | 1 | 2 | 0 | 6 | Medium |
| Deng et al,[27] 2020 | 1 | 1 | 0 | 1 | 1 | 2 | 0 | 6 | Medium |
| Dong et al,[28] 2020 | 1 | 1 | 0 | 1 | 2 | 2 | 0 | 7 | High |
| He et al,[29] 2020 | 1 | 1 | 1 | 1 | 1 | 1 | 1 | 7 | High |
| Lee et al,[30] 2020 | 1 | 1 | 0 | 0 | 2 | 1 | 1 | 6 | Medium |

| Study | Was the follow up > 3 years? (Yes=1, No/NR=0) | Was the sample size of training dataset at least 100? (Yes=1, No/NR=0) | Were there appropriate methods of imputation used for missing data? (Yes=1, No/NR=0) | Were methylation biomarkers selected or assessed by adjusting for age, sex, and tumor stage? (Yes=1, No/NR=0) | Was validation performed? (No =0, Internal =1, External =2) | Were relevant performance measures evaluated appropriately (No=0, only discrimination=1, both discrimination and calibration=2) | Was Cross-validation or Bootstrapping used to evaluate performance (Yes =1, No/NR=0) | Total score | Quality |
|---|---|---|---|---|---|---|---|---|---|
| Li, C et al,[31] 2020 | 1 | 1 | 1 | 1 | 1 | 1 | 0 | 6 | Medium |
| Li, T et al,[32] 2020 | 1 | 1 | 0 | 1 | 1 | 1 | 0 | 5 | Medium |
| Lian, et al,[33] 2020 | 1 | 1 | 1 | 1 | 1 | 1 | 1 | 7 | High |
| Luo, H et al,[34] 2020 | 1 | 1 | 0 | 1 | 1 | 1 | 0 | 5 | Medium |
| Luo, R et al,[35] 2020 | 1 | 1 | 0 | 1 | 2 | 1 | 0 | 6 | Medium |
| Lv et al,[36] 2020 | 1 | 1 | 1 | 0 | 1 | 1 | 1 | 6 | Medium |
| Ma, H et al,[37] 2020 | 1 | 0 | 1 | 0 | 2 | 1 | 0 | 5 | Medium |
| Ma, X et al,[38] 2020 | 1 | 1 | 1 | 1 | 1 | 2 | 0 | 7 | High |
| Macías-García et al,[39] 2020 | 1 | 0 | 0 | 0 | 1 | 1 | 0 | 3 | Low |
| Shi et al,[40] 2020 | 1 | 1 | 1 | 0 | 1 | 1 | 0 | 5 | Medium |
| Tian et al,[41] 2020 | 1 | 1 | 1 | 1 | 1 | 2 | 0 | 7 | High |
| Wang, X et al,[42] 2020 | 1 | 1 | 0 | 1 | 1 | 2 | 1 | 7 | High |
| Wang, Y et al,[43] 2020 | 1 | 1 | 1 | 1 | 1 | 1 | 0 | 6 | Medium |
| Xiang et al,[44] 2020 | 1 | 1 | 0 | 1 | 2 | 2 | 0 | 7 | High |
| Yu et al,[45] 2020 | 1 | 0 | 1 | 0 | 1 | 1 | 1 | 5 | Medium |
| Zhang, C et al,[46] 2020 | 1 | 1 | 0 | 0 | 2 | 2 | 0 | 6 | Medium |
| Zhang, E et al,[47] 2020 | 1 | 1 | 0 | 0 | 1 | 1 | 0 | 4 | Medium |
| Zheng et al,[48] 2020 | 1 | 1 | 0 | 0 | 0 | 0 | 0 | 2 | Low |
| Bedon et al,[49] 2021 | 1 | 1 | 0 | 0 | 1 | 1 | 1 | 5 | Medium |
| Chen et al,[50] 2021 | 1 | 0 | 1 | 0 | 1 | 1 | 0 | 4 | Medium |
| Hao et al,[51] 2021 | 1 | 1 | 1 | 1 | 0 | 1 | 0 | 5 | Medium |
| Huang, G et al,[52] 2021 | 1 | 1 | 1 | 1 | 0 | 1 | 0 | 5 | Medium |
| Huang, GJ et al,[53] 2021 | 1 | 1 | 1 | 0 | 2 | 1 | 1 | 7 | High |
| Huang, H et al,[54] 2021 | 1 | 1 | 0 | 1 | 2 | 2 | 0 | 7 | High |
| Huang, W et al,[55] 2021 | 1 | 1 | 0 | 1 | 2 | 2 | 0 | 7 | High |
| Lee et al,[56] 2021 | 1 | 1 | 0 | 1 | 2 | 1 | 0 | 6 | Medium |
| Li, K et al,[57] 2021 | 1 | 1 | 1 | 1 | 0 | 1 | 0 | 5 | Medium |
| Li, X et al,[58] 2021 | 1 | 1 | 0 | 1 | 1 | 1 | 0 | 5 | Medium |
| Peng et al,[59] 2021 | 1 | 1 | 0 | 1 | 1 | 1 | 0 | 5 | Medium |
| Shang et al,[60] 2021 | 0 | 0 | 0 | 0 | 0 | 1 | 1 | 2 | Low |
| Sun et al,[61] 2021 | 1 | 1 | 0 | 1 | 1 | 1 | 0 | 5 | Medium |

| Study | Was the follow up > 3 years? (Yes=1, No/NR=0) | Was the sample size of training dataset at least 100? (Yes=1, No/NR=0) | Were there appropriate methods of imputation used for missing data? (Yes=1, No/NR=0) | Were methylation biomarkers selected or assessed by adjusting for age, sex, and tumor stage? (Yes=1, No/NR=0) | Was validation performed? (No =0, Internal =1, External =2) | Were relevant performance measures evaluated appropriately (No=0, only discrimination=1, both discrimination and calibration=2) | Was Cross-validation or Bootstrapping used to evaluate performance (Yes =1, No/NR=0) | Total score | Quality |
|---|---|---|---|---|---|---|---|---|---|
| Tan et al,[62] 2021 | 1 | 1 | 0 | 1 | 2 | 1 | 0 | 6 | Medium |
| Wang, L et al,[63] 2021 | 1 | 1 | 1 | 1 | 1 | 1 | 0 | 6 | Medium |
| Wang, T et al,[64] 2021 | 1 | 1 | 0 | 1 | 0 | 1 | 1 | 5 | Medium |
| Wu et al,[65] 2021 | 1 | 1 | 1 | 1 | 1 | 1 | 0 | 6 | Medium |
| Xu et al,[66] 2021 | 1 | 1 | 0 | 1 | 2 | 2 | 0 | 7 | High |
| Yin L et al,[67] 2021 | 1 | 1 | 1 | 1 | 1 | 1 | 0 | 6 | Medium |
| Yin X et al,[68] 2021 | 1 | 1 | 1 | 1 | 1 | 2 | 0 | 7 | High |
| Yu et al,[69] 2021 | 1 | 1 | 1 | 0 | 2 | 1 | 1 | 7 | High |

**Supplementary Table 5: Studies using clustering-based pipelines**

| Author, year | Pre-sreening CpGs | Clustering samples | Finding intracluster-specific CPGs/MDGs | Training model (further feature selection) | Validation |
|---|---|---|---|---|---|
| Wei et al,[1] 2002 | dmCpGs selected comparing TT/NT | Hierarchical clustering | Batch sequential forward selection | | |
| Walker et al,[2] 2011 | Not performed | Hierarchical clustering | dmCpGs across clusters | | |
| Sigalotti et al,[3] 2012 | Not performed | K-means clustering | Nearest shrunken centroid | | |
| Chambwe et al,[4] 2014 | Not performed | Hierarchical clustering, Consensus clustering | dmCpGs across clusters | | |
| Mah et al,[6] 2014 | CpGs with high variance | Consensus clustering | dmCpGs across clusters | | |
| Wang et al,[7] 2014 | CpGs associated with prognosis by univariable and multivariable analysis | Recursive partitioning mixture model | Not performed | Bayesian network classifier | Sample-Splitting |
| Ouchi et al,[8] 2015 | CpGs with high variance | Hierarchical clustering | Random forest | | Sample-Splitting |
| Saito et al,[14] 2016 | CpGs with high variance | Hierarchical clustering, Consensus clustering | dmCpGs across clusters | | |
| Stieglitz et al,[17] 2017 | CpGs with high variance | Hierarchical clustering | Other | | |
| Zhang et al,[18] 2018 | CpGs associated with prognosis by univariable and multivariable analysis | Consensus clustering | dmCpGs across clusters | Bayesian network classifier | Cross-validation External validation |
| Chen et al,[19] 2019 | CpGs associated with prognosis by univariable and multivariable analysis | Consensus clustering | dmCpGs across clusters | Bayesian network classifier | Cross-validation Sample-Splitting |
| Li et al,[21] 2019 | CpGs with high variance | Consensus clustering | dmCpGs across clusters | | |
| Yang et al,[23] 2019 | CpGs associated with prognosis by univariable and multivariable analysis | Consensus clustering | dmCpGs in a seed cluster | Cox | Sample-Splitting |
| Bian et al,[24] 2020 | CpGs associated with prognosis by univariable and multivariable analysis | Consensus clustering | dmCpGs across clusters | LASSO-stepwise AIC-Cox | Sample-Splitting |
| Cai et al,[25] 2020 | CpGs associated with prognosis by univariable and multivariable analysis | Consensus clustering | dmCpGs across clusters | Stepwise-Cox | Sample-Splitting |
| He et al,[29] 2020 | CpGs associated with prognosis by univariable and multivariable analysis | Consensus clustering | dmCpGs across clusters | Bayesian network classifier | Cross-validation Sample-Splitting |
| Li, C et al,[31] 2020 | CpGs associated with prognosis by univariable and multivariable analysis | Consensus clustering | dmCpGs across clusters | LASSO-Cox | Sample-Splitting |
| Li, T et al,[32] 2020 | CpGs associated with prognosis by univariable and multivariable analysis | Consensus clustering | dmCpGs in a seed cluster | Cox | Sample-Splitting |
| Lian, et al,[33] 2020 | CpGs associated with prognosis by univariable and multivariable analysis | Consensus clustering | dmCpGs across clusters | Random forest classifier | Sample-Splitting Cross validation |
| Ma, H et al,[37] 2020 | CpGs associated with prognosis by univariable and multivariable analysis | Consensus clustering | Hub CpGs obtained by WGCNA and a seed cluster | | Sample-Splitting External validation |
| Shi et al,[40] 2020 | CpGs associated with prognosis by univariable and multivariable analysis | Consensus clustering | dmCpGs in a seed cluster | Cox | Sample-Splitting |

| Author, year | Pre-sreening CpGs | Clustering samples | Finding intracluster-specific CPGs/MDGs | Training model (further feature selection) | Validation |
|---|---|---|---|---|---|
| Tian et al,[41] 2020 | CpGs associated with prognosis by univariable and multivariable analysis | Consensus clustering | dmCpGs in a seed cluster | Cox | Sample-Splitting |
| Wang, X et al,[42] 2020 | CpGs associated with prognosis by univariable and multivariable analysis | Hierarchical clustering | dmCpGs across clusters | LASSO-Cox | Sample-Splitting<br>Bootstrapping |
| Wang, Y et al,[43] 2020 | CpGs with high variance<br>CpGs associated with prognosis by univariable and multivariable analysis | Hierarchical clustering | Hub CpGs obtained by WGCNA and a seed cluster | Multifactor regression | Sample-Splitting |
| Xiang et al,[44] 2020 | CpGs associated with prognosis by univariable and multivariable analysis | Consensus clustering | dmCpGs across clusters | LASSO-Cox | External validation |
| Zheng et al,[48] 2020 | Not performed | Consensus clustering | dmCpGs across clusters | | |
| Chen et al,[50] 2021 | CpGs associated with prognosis by univariable and multivariable analysis | Consensus clustering | Hub CpGs obtained by WGCNA and a seed cluster | | Sample-Splitting |
| Huang, G et al,[52] 2021 | CpGs associated with prognosis by univariable and multivariable analysis | Consensus clustering | dmCpGs in a seed cluster | AIC-Cox | |
| Li, K et al,[57] 2021 | CpGs associated with prognosis by univariable and multivariable analysis | Consensus clustering | dmCpGs in a seed cluster | LASSO-Cox | |
| Li, X et al,[58] 2021 | CpGs associated with prognosis by univariable and multivariable analysis | Consensus clustering | dmCpGs in a seed cluster | Cox | Sample-Splitting |
| Wang et al,[63] 2021 | CpGs associated with prognosis by univariable and multivariable analysis | Consensus clustering | dmCpGs in a seed cluster | Cox | Sample-Splitting |
| Wu et al,[65] 2021 | CpGs associated with prognosis by univariable and multivariable analysis | Consensus clustering | dmCpGs in a seed cluster | AIC-Cox | Sample-Splitting |
| Xu et al,[66] 2021 | CpGs associated with prognosis by univariable and multivariable analysis | Consensus clustering | dmCpGs in a seed cluster | LASSO-Cox | Sample-Splitting<br>External validation |
| Yin, L,[67] 2021 | CpGs associated with prognosis by univariable and multivariable analysis | Consensus clustering | Hub CpGs obtained by WGCNA and a seed cluster | | |
| Yin, X,[68] 2021 | CpGs associated with prognosis by univariable and multivariable analysis | Consensus clustering | dmCpGs in a seed cluster | LASSO-Cox | Sample-Splitting |

**Supplementary Table 6: Studies using supervised feature selection-based pipelines**

| Author, year | Pre-screening CpGs | Initial feature selection | Training model (feature selection) | Validation |
|---|---|---|---|---|
| Fleischer et al,[5] 2014 | dmCpGs selected comparing TT/NT<br>dmCpGs correlated with gene expression | LASSO-Cox | Cox | External validation |
| Wei et al,[10] 2015 | dmCpGs selected comparing TT/NT | LASSO-Logistic | LASSO-Cox | External validation |
| Villanueva et al,[9] 2015 | dmCpGs selected comparing TT/NT | Not performed | Random survival forest | Bootstrapping |
| Bjaanaes et al,[11] 2016 | CpGs correlated with gene expression<br>CpGs with high variance | LASSO-Cox | Cox | External validation |
| Lim et al,[12] 2016 | Not performed | Centroid feature selection, SVM-RFE | | |
| Lu et al,[13] 2016 | Not performed | Univariable-Logistic | LASSO-Logistic | Cross-validation<br>Bootstrapping trai |
| Hao et al,[15] 2017 | CpGs associated with prognosis by univariable analysis | LASSO-Cox, Boosting | Cox | Sample-Splitting |
| Qiu et al,[16] 2017 | A threshold value of 0.15 for the average b in each group | LASSO-Logistic<br>SVM-RFE | Lasso Cox | Sample-Splitting,<br>External validation |
| Dong et al,[20] 2019 | dmCpGs selected comparing TT/NT<br>CpGs associated with prognosis by univariable analysis | SVM-RFE | FW-SVM | Sample-Splitting,<br>Cross validation,<br>External validation |
| Wang et al,[22] 2019 | CpGs with high variance | LASSO-Logistic<br>RFE | Cox | Cross validation |
| Ma, X et al,[38] 2020 | dmCpGs selected comparing early/advanced tumor | LASSO-Cox | Multivariable Cox with stepwise selection | Sample-Splitting |
| Chen et al,[26] 2020 | dmCpGs selected comparing early and advanced tumor<br>dmCpGs associated with prognosis by univariable analysis | LASSO-Cox | Multivariable Cox | Sample-Splitting |
| Deng et al,[27] 2020 | dmCpGs selected comparing TT/NT<br>dmCpGs associated with prognosis by univariable analysis | LASSO-Logistic<br>SVM-RFE | LASSO-Cox | Sample-Splitting |
| Dong et al,[28] 2020 | CpGs associated with prognosis by univariable analysis | Random survival forests-variable hunting | Multivariable Cox | Sample-Splitting,<br>External validation |
| Luo, H et al,[34] 2020 | CpGs associated with prognosis by univariable analysis | LASSO-Cox | Cox | Sample-Splitting |
| Luo, R et al,[35] 2020 | Differential CpGs correlated with gene expression | LASSO-Logistic<br>Random forest | LASSO Cox<br>Univariable Cox | Sample-Splitting,<br>External validation |
| Zhang, C et al,[46] 2020 | MDG selected by comparing TT/NT and by intersecting between DEG and DMG<br>MDG associated with prognosis by univariable analysis | LASSO-Cox | Cox | External validation |

| Author, year | Pre-screening CpGs | Initial feature selection | Training model (feature selection) | Validation |
|---|---|---|---|---|
| Zhang, E et al,[47] 2020 | dmCpGs selected comparing TT/NT | LASSO-Cox | Cox | Sample-Splitting |
| Bedon et al,[49] 2021 | CpGs with high variance | Random forest-Recursive Features Selection<br>Random forest - All relevant features analysis (Boruta) | Univariable Cox<br>Multivariable Cox with stepwise feature selection | Sample-Splitting,<br>Cross validation,<br>Bootstrapping |
| Huang, H et al,[54] 2021 | DMG selected by comparing TT/NT | LASSO-Cox | Multivariable Cox | External validation |
| Huang, W et al,[55] 2021 | DMG selected by comparing TT/NT and by intersecting between DMG, genes associated with prognosis, and genes related with copy number variance | LASSO-Cox | Cox | External validation |
| Hao et al,[51] 2021 | CpGs associated with prognosis by univariable analysis | LASSO-Cox | Cox | Random selection from the training set |
| Lee et al,[56] 2021 | MDGs selected by comparing TT/NT and by intersecting between DEG and DMG<br>MDG associated with survival by univariable Cox | Random survival forests-variable hunting | Multivariable Cox | Sample-Splitting,<br>External validation |
| Peng et al,[59] 2021 | MDGs selected by comparing TT/NT and are correlated with gene expression | LASSO-Cox | Cox | Sample-Splitting |
| Shang et al,[60] 2021 | CpGs associated with prognosis by univariable Logistic analysis | LASSO-Logistic<br>Random forest<br>Support vector machine | LASSO-Logistic | Cross-validation,<br>External validation |
| Sun et al,[61] 2021 | DMG selected by comparing TT/NT and by intersecting between DEG<br>WGCNA co-methylation analysis | Univariable Cox<br>Multivariable Cox | LASSO-Cox | Sample-Splitting |
| Tan et al,[62] 2021 | DMG selected by comparing TT/NT and by intersecting between DEG | Multivariable Cox | LASSO-Cox | External validation |
| Yu et al,[69] 2021 | Identifying methylation-correlated blocks | LASSO-Cox | Elastic-net<br>Support vector machine<br>Mutlivariable Cox<br>Stacking ensemble model | Cross-validation,<br>External validation |

**Supplementary Table 7: Studies using deep learning-based pipelines**

| ***Only DNA methylation biomarkers*** | | | | | | |
|---|---|---|---|---|---|---|
| **Author, year** | **Feature transformation** | **Select survival associated nodes** | **CpGs weight calculation** | **Finding the best MDGs** | | |
| Macías-García et al,[39] 2020 | Autoencoder | Univariable Cox | Calculate CpGs weight in every significant node, and then sum the weight of CpGs corresponding to the same gene | Intersection between the best genes of the significant nodes | | |
| ***Multi-omics*** | | | | | | |
| **Author, year** | **Feature transformation** | **Selecting survival associated nodes** | **Clutering samples** | **Finding intracluster-specific MDGs** | **Training model** | **Validation** |
| Lee et al,[30] 2020 | Autoencoder | Univariable Cox LASSO-Cox | K-means clustering | dmMDGs across clusters | Random forest | Cross validation, External validation |
| Lv et al,[36] 2020 | Autoencoder | Univariable Cox | K-means clustering | dmMDGs across clusters | Support vector machine | Cross validation |
| Yu et al, 2020 | Autoencoder | Univariable Cox | K-means clustering | dmMDGs across clusters | Support vector machine | Cross validation, External validation |
| Huang, GJ et al,[53] 2021 | Bidirectional deep neural networks | Univariable Cox | K-means clustering | dmMDGs across clusters | Support vector machine | Cross validation, External validation |
| Wang, T et al,[53] 2021 | Autoencoder | Univariable Cox | K-means clustering | dmMDGs across clusters | Support vector machine | Cross-validation |

**Supplementary Table 8: Performance measurement of identified DNA methylation biomarkers**

| Author, year | Evaluation object | Discrimination | | | Calibration |
|---|---|---|---|---|---|
| | | Cox regression analysis | C-index | ROC/AUC | |
| ***Clustering-based*** | | | | | |
| Wei et al,[1] 2002 | Clusters | The BSFS analysis revealed a subset of 182 hypermethylated loci for the two groups, showing definitively that more loci were hypermethylated per tumor in group 1 than in group 2. This differential methylation in ovarian tumors correlated significantly with PFS (p< 0.001, log-rank test). | | | |
| Mah et al,[6] 2014 | Clusters | cHR (95%CI), training set, OS: 4.73 (1.31-16.91)<br>cHR (95%CI), training set, PFS: 3.99 (1.61-9.86) | | | |
| Wang et al,[7] 2014 | Clusters | aHR (95%CI), sample-splitting: 1.92 (1.25-2.94) | | | |
| Li et al,[21] 2019 | Clusters | Whole dataset, Log-rank test: p=0.03 | | | |
| Yang et al,[23] 2019 | Prognostic index | Sample-splitting, Log-rank test: p = 0.032 | | | |
| Bian et al,[24] 2020 | Prognostic index | Sample-splitting, aHR (95%CI), 2.12 (1.19-3.78) | | Sample-splitting, 3-year: 0.70 | |
| Cai et al,[24] 2020 | Prognostic index | Sample-splitting, Log-rank test: p < 0.05 | Prognostic model including other clinical variables, sample splitting: 0.76 | Sample-splitting, DNA methylation signatures: 0.5-year: 0.75; 1-year: 0.83; 3-year: 0.72<br>Prognostic model including other clinical variables, 3-year: 0.80 | Prognostic model including other clinical variables, sample splitting: The calibration curve for the probability of survival for 3 years in the test set also showed good agreement between prediction and observation |
| He et al,[29] 2020 | Clusters | Sample-splitting, Log-rank test: p < 0.001 | | | |
| Li, C et al,[31] 2020 | Prognostic index | Sample-splitting, Log-rank test: p =0.04 | | | |
| Li, T et al,[32] 2020 | Clusters | Sample-splitting, From the survivorship curve, we could see that patients in high-risk group had lower survival probability than in low-risk group | | | |
| Lian et al,[33] 2020 | Clusters | Sample-splitting, Log-rank test: p =0.026 | | | |
| Ma, H et al,[37] 2020 | Clusters | External validation, Log-rank test: p = <0.001 | | | |
| Shi et al,[40] 2020 | Prognostic index | Sample-splitting, Log-rank test: p = 0.02 | | Training set, DNA methylation signature: 0.78 | |
| Tian et al,[41] 2020 | Prognostic index | Training set aHR (95%CI), 2.81 (1.80-4.40) | Prognostic model including other clinical variables, training set: 0.77 | Training set, DNA methylation signature: 0.69 | Prognostic model including other clinical variables: calibration plot chart showed that the actual survival rate of 5 years was in good agreement with the predicted survival rate |
| Wang, X et al,[42] 2020 | Prognostic index | Sample-splitting, cHR (95%CI), OS, 2.80 (1.17-6.68)<br>PFS, 2.31 (1.08-4.97) | Sample-splitting, OS: 0.72 (0.60-0.83) | Sample-splitting, DNA methylation signatures: 3-year: 0.68 (0.58-0.78); 5-year: 0.70 (0.60-0.81) | The calibration curves displayed good agreement between prediction and actual observation in the 3-year and 5-year OS probability |
| Wang, Y et al,[43] 2020 | Prognostic index | Sample-splitting, Log-rank test: p < 0.001 | | | |
| Zheng et al,[48] 2020 | Prognostic index | | | Whole dataset, DNA methylation signature, 3-year: 0.76, 5-year: 0.81, 7-year: 0.83 | |

| Author, year | Evaluation object | Discrimination | | | Calibration |
|---|---|---|---|---|---|
| | | Cox regression analysis | C-index | ROC/AUC | |
| ***Clustering-based*** | | | | | |
| Chen et al,[50] 2021 | Clusters | Sample-splitting, Log-rank test: p = 0.02 | | | |
| Huang, G et al,[52] 2021 | | Whole dataset, Log-rank test: p = 0.001 | | Whole dataset, 5-year: 0.657 | |
| Li, K et al,[57] 2021 | Prognostic index | Whole dataset, aHR (95%CI), 1.25 (1.19-1.31) | | Whole dataset, DNA methylation signature, 1-year: 0.80, 3-year: 0.79, 5-year: 0.79 | |
| Li, X et al,[58] 2021 | Prognostic index | Whole dataset, aHR (95%CI), 2.67 (2.10-3.39) | | Sample-splitting, 1-year: 0.54, 3-year: 0.57, 5-year: 0.66 | |
| Wang, L et al,[63] 2021 | Prognostic index | Sample-splitting, Log-rank test: p = 0.04 | | Sample-splitting: 0.77 | |
| Wu et al,[65] 2021 | Prognostic index | Sample-splitting, Log-rank test: p = 0.03 | | Sample-splitting: 0.60 | |
| Xu et al,[66] 2021 | Prognostic index | Sample-splitting, aHR (95%CI), 1.18 (1.05-1.32) | | External validation: 0.68 | Prognostic model including other clinical variables: Calibrated curves demonstrated good prognosis prediction accuracy of one-, two-, and three-year overall survival in the as-constructed nomogram |
| Xiang et al,[44] 2020 | Prognostic index | Sample-splitting, Log-rank test: p = 0.04 | Sample-splitting, Stomach: 0.66; Colon: 0.68 | Whole dataset, prognostic model including other clinical variable, Stomach, 3-year: 0.79, 5-year: 0.778; Colon, 3-year: 0.91, 5-year: 0.86 | The calibration curves for predicting 3-year and 5-year OS are in good agreement with the actual observations |
| Yin, L et al,[67] 2021 | Clusters | Sample-splitting, Log-rank test: p = 0.02 | | | |
| Yin, X et al,[68] 2021 | Prognostic index | Whole dataset, aHR (95%CI), 8.11 (3.67-17.92) | DNA methylation signatures: 0.64; Prognostic model including other clinical variables: 0.66 | Sample-splitting, DNA methylation signatures: 1-year: 0.72; 3-year: 0.86; 5-year: 0.75<br>Prognostic model including other clinical variables: 1-year: 0.70; 3-year: 0.83; 5-year: 0.77 | The calibration curves of the risk score and nomogram model showed that the two models present satisfied coherence between the actual survival and predicted survival rates. |
| **Supervised feature selection-based** | | | | | |
| Fleischer et al,[5] 2014 | Prognostic index | aHR (95%CI), external validation: 2.09 (1.21-3.59) | | | |
| Villanueva et al,[9] 2015 | Prognostic index | aHR (95%CI), training set, OS: 13.35 (7.94-22.42); RFS: 5.8 (3.1-11)<br>External validation: RFS, Log-rank test: p = 0.01 | | | |
| Wei et al,[10] 2015 | Prognostic index | aHR (95%CI), external validation1: 3.73 (2.28-6.09); external validation2: 3.36 (1.78-6.34); external validation3: 1.80 (1.11-2.93) | | | |
| Bjaanaes et al,[11] 2016 | Prognostic index | aHR (95%CI), external validation: 1.61 (0.92-2.45) | | | |
| Lim et al,[12] 2016 | Prognostic index | | Training set: 0.52-0.58 | | |
| Hao et al,[15] 2017 | Prognostic index | | Sample-splitting, Breast: 0.63, Lung: 0.64 | | |

| Author, year | Evaluation object | Discrimination | | | Calibration |
|---|---|---|---|---|---|
| | | Cox regression analysis | C-index | ROC/AUC | |
| **Supervised feature selection-based** | | | | | |
| Qiu et al,[16] 2017 | Prognostic index | RFS, aHR (95%CI), external validation1: 2.83 (1.89-4.23); external validation2: 2.42 (1.37-4.20)<br>OS, aHR (95%CI), external validation1: 2.81 (1.12-4.41); external validation2: 2.91 (1.48-5.71) | Prognostic model including other clinical variables: external validation1: 0.697; external validation2: 0.693 | Only DNA methylation signatures,<br>2-year: 0.71 (0.67-0.74),<br>5-year: 0.72 (0.68-0.75)<br>DNA methylation +clinical variables,<br>2-year: 0.79 (0.75-0.81),<br>5-year: 0.79 (0.76-0.82) | Prognostic model including other clinical variables: the calibration plots for the 5-year RFS rate were predicted well in the three validation cohorts |
| Dong et al,[20] 2019 | Prognostic index | Sample-splitting: cHR (95%CI), High-risk vs. low-risk: cHR = 8.95 (1.96-40.92)<br>High- risk vs. medium-risk: cHR = 3.12 (1.31-7.46)<br>High-risk vs. (medium-risk + low-risk):4.77 (2.09-10.90)<br>High-risk + medium-risk) vs. low-risk: 6.41 (1.49-27.54) | | | |
| Wang et al,[22] 2019 | Prognostic index | Training set, Log-rank test: p < 0.0001 | | Training set, DNA methylation signature: 500-day: 0.69; 1000-day: 0.68; 1500-day: 0.70; 2000-day:0.69; 2500-day:0.74; 3000-day:0.76.<br>Prognostic model including other clinical variables: 500-day: 0.72; 1000-day: 0.68; 1500-day: 0.70; 2000-day:0.69; 2500-day:0.72; 3000-day:0.70 | Training set, prognostic model including other clinical variables: In calibration plot, the predicted OS of LUAD patients by the nomogram was highly consistent with the observed 3- and 5-year OS of lung patients |
| Chen et al,[26] 2020 | Prognostic index | Sample-splitting, DNA methylation signatures: aHR (95%CI), 1.79 (1.09-2.94) | Prognostic model including other clinical variables: 0.8 (0.70-0.89) | Sample-splitting, DNA methylation signatures: 1-year: 0.91; 3-year: 0.80; 5-year: 0.82. Prognostic model including other clinical variables: 1-year: 0.85; 3-year: 0.78, 5-year: 0.80 | Prognostic model including other clinical variables: Calibration plot yielded a high predictive value |
| Dong et al,[28] 2020 | Prognostic index | Sample-splitting, DNA methylation signatures: cHR (95%CI), 1.14 (1.02-1.27) | | DNA methylation signatures: Sample-splitting, 0.72; External validation: 0.74 | In the training group, the calibration chart of the 5-year operating system was well-predicted |
| Deng et al,[27] 2020 | Prognostic index | Sample-splitting, cHR (95%CI), 2.17 (1.23-3.85) | Prognostic model including other clinical variables, sample splitting: 0.69 | Sample-splitting, DNA methylation signatures: 3-year: 0.65; 5-year: 0.64<br>Prognostic model including other clinical variables: 3-year: 0.84; 5-year: 0.74 | The calibration curve of the nomogram showed good agreement between prediction and observation in the test set |
| Luo, H et al,[34] 2020 | Prognostic index | Sample-splitting, DNA methylation signatures: aHR (95%CI), 1.63 (1.03-2.57) | Prognostic model including other clinical variables, sample splitting: 0.78 | Sample-splitting, DNA methylation signatures, 6-month: 0.72 (0.63-0.81); Prognostic model including other clinical variables: 6-month: 0.87 (0.82-0.93) | Prognostic model including other clinical variables: the probability of 3-year overall survival as predicted by the nomogram was close to the observed survival |
| Luo, R et al,[35] 2020 | Prognostic index | Training set aHR (95%CI), RFS: 1.38 (1.07-1.79); OS: 1.4 (1.11-1.77) | | DNA methylation signatures external validation, 2-year, RSF: 0.82; | |
| Ma, X et al,[38] 2020 | Prognostic index | Sample-splitting, Log-rank test: p < 0.001<br>whole dataset, aHR (95%CI), 2.55 (1.99-3.27) | Prognostic model including other clinical variables, whole dataset: 0.76 | DNA methylation signatures: Sample-splitting, 5-year 0.74<br>Prognostic model including other clinical variables, whole dataset, 1-year: 0.81, 3-year: 0.79, 5-year: 0.81 | Prognostic model including other clinical variables: calibration plot yielded a high value |

| Author, year | Evaluation object | Discrimination | | | Calibration |
|---|---|---|---|---|---|
| | | Cox regression analysis | C-index | ROC/AUC | |
| **Supervised feature selection-based** | | | | | |
| Zhang, C et al,[46] 2020 | Prognostic index | External validation, Log-rank test: p = 0.03<br>Whole dataset, aHR (95%CI), 4.19 (1.68-8.46) | | External validation, DNA methylation signature: 0.88 | Prognostic model including other clinical variables: calibration plot chart showed the consistency between the 5-year survival predicted by the nomogram survival model and the actual 5-year survival. |
| Zhang, E et al,[47] 2020 | Prognostic index | Whole dataset, aHR (95%CI), 10.3 (2.43-43.30) | | Whole dataset, DNA methylation signature, 3-year: 0.76, 5-year: 0.81, 7-year: 0.83 | |
| Bedon et al,[49] 2021 | Prognostic index | Sample-splitting, Log-rank test: p = 0.04 | | | |
| Hao et al,[51] 2021 | Prognostic index | Whole dataset, aHR (95%CI), 2.54 (1.49-4.35) | | Sample-splitting, 1-year: 0.65, 3-year: 0.77, 5-year: 0.82 | |
| Huang, H et al,[54] 2021 | Prognostic index | External validation, aHR (95%CI), 1.44 (1.05-1.96) | Prognostic model including other clinical variables, external validation: 0.71 | External validation, DNA methylation signature, 3-year: 0.70, 5-year: 0.72, 8-year: 0.80 | In the training set, calibration plots demonstrated that the nomogram had similar predictive performance compared with an ideal model in predicting the 5- year OS for CRC patients. Similar results were observed in the testing set. |
| Huang, W et al,[55] 2021 | Prognostic index | Wole dataset, aHR (95%CI), 10.38 (5.14–20.96) | Prognostic model including other clinical variables, training, 1-year: 0.717, 3-year: 0.714, 5-year: 0.715 | DNA methylation signature, external validation1, 1-year: 0.677, 3-year: 0.675, 5-year: 065; external validation2, 1-year: 0.521, 3-year: 0.781, 5-year: 0.758 | Prognostic model including other clinical variables: the results of the calibration chart indicate good agreement between the predicted and actual observations |
| Lee et al,[56] 2021 | Prognostic index | Whole dataset, aHR (95%CI), 2.88 (1.88-4.2) | | DNA methylation signature, external validation: 0.74 | |
| Peng et al,[59] 2021 | Prognostic index | Whole dataset, aHR (95%CI), 1.60 (1.28-2.01) | | Whole dataset: 0.68 | |
| Shang et al,[60] 2021 | Prognostic index | | | Training set: 0.97 | |
| Sun et al,[61] 2021 | Prognostic index | Sample-splitting, aHR (95%CI), 2.87 (1.15-7.12) | DNA methylation signatures only, Whole dataset, 0.72 | Sample-splitting, DNA methylation signatures: 0.82 | |
| Tan et al,[62] 2021 | Prognostic index | External validation, aHR (95%CI), 1.75 (1.15-2,70) | | | |
| Yu et al,[69] 2021 | Prognostic index | Whole dataset, aHR (95%CI), 2.28 (1.19–4.37) | | DNA methylation signature, 5-year cross-validation: 0.62, sample-splitting: 0.70 | |
| ***Deep learning-based*** | | | | | |
| Wang, T et al,[64] 2021 | Prognostic index | | Training set, multi-omics panel only: 0.694; combined with clinical information: 0.71 | Training set, multi-omics panel only: 0.742; combined with clinical information: 0.789 | |

Multivariate analysis and external validation were prioritized over univariable analysis and internal validation.

### References

1. Wei SH, Chen CM, Strathdee G, et al. Methylation microarray analysis of late-stage ovarian carcinomas distinguishes progression-free survival in patients and identifies candidate epigenetic markers. *Clin Cancer Res* 2002; **8**(7): 2246-52.
2. Walker BA, Wardell CP, Chiecchio L, et al. Aberrant global methylation patterns affect the molecular pathogenesis and prognosis of multiple myeloma. *Blood* 2011; **117**(2): 553-62.
3. Sigalotti L, Covre A, Fratta E, et al. Whole genome methylation profiles as independent markers of survival in stage IIIC melanoma patients. *J Transl Med* 2012; **10**: 185.
4. Chambwe N, Kormaksson M, Geng H, et al. Variability in DNA methylation defines novel epigenetic subgroups of DLBCL associated with different clinical outcomes. *Blood* 2014; **123**(11): 1699-708.
5. Fleischer T, Frigessi A, Johnson KC, et al. Genome-wide DNA methylation profiles in progression to in situ and invasive carcinoma of the breast with impact on gene transcription and prognosis. *Genome Biology* 2014; **15**(8).
6. Mah WC, Thurnherr T, Chow PK, et al. Methylation profiles reveal distinct subgroup of hepatocellular carcinoma patients with poor prognosis. *PLoS One* 2014; **9**(8): e104158.
7. Wang C, Cicek MS, Charbonneau B, et al. Tumor hypomethylation at 6p21.3 associates with longer time to recurrence of high-grade serous epithelial ovarian cancer. *Cancer Res* 2014; **74**(11): 3084-91.
8. Ouchi K, Takahashi S, Yamada Y, et al. DNA methylation status as a biomarker of anti-epidermal growth factor receptor treatment for metastatic colorectal cancer. *Cancer Sci* 2015; **106**(12): 1722-9.
9. Villanueva A, Portela A, Sayols S, et al. DNA methylation-based prognosis and epidrivers in hepatocellular carcinoma. *Hepatology* 2015; **61**(6): 1945-56.
10. Wei JH, Haddad A, Wu KJ, et al. A CpG-methylation-based assay to predict survival in clear cell renal cell carcinoma. *Nature Communications* 2015; **6**.
11. Bjaanaes MM, Fleischer T, Halvorsen AR, et al. Genome-wide DNA methylation analyses in lung adenocarcinomas: Association with EGFR, KRAS and TP53 mutation status, gene expression and prognosis. *Molecular Oncology* 2016; **10**(2): 330-43.
12. Lim AM, Wong NC, Pidsley R, et al. Genome-scale methylation assessment did not identify prognostic biomarkers in oral tongue carcinomas. *Clinical Epigenetics* 2016; **8**(1).
13. Lu J, Cowperthwaite MC, Burnett MG, Shpak M. Molecular Predictors of Long-Term Survival in Glioblastoma Multiforme Patients. *PLoS One* 2016; **11**(4): e0154313.
14. Saito Y, Nagae G, Motoi N, et al. Prognostic significance of CpG island methylator phenotype in surgically resected small cell lung carcinoma. *Cancer Sci* 2016; **107**(3): 320-5.
15. Hao X, Luo H, Krawczyk M, et al. DNA methylation markers for diagnosis and prognosis of common cancers. *Proceedings of the National Academy of Sciences of the United States of America* 2017; **114**(28): 7414-9.
16. Qiu J, Peng B, Tang Y, et al. CpG Methylation Signature Predicts Recurrence in Early-Stage Hepatocellular Carcinoma: Results From a Multicenter Study. *J Clin Oncol* 2017; **35**(7): 734-42.
17. Stieglitz E, Mazor T, Olshen AB, et al. Genome-wide DNA methylation is predictive of outcome in juvenile myelomonocytic leukemia. *Nat Commun* 2017; **8**(1): 2127.
18. Zhang S, Wang Y, Gu Y, et al. Specific breast cancer prognosis-subtype distinctions based on DNA methylation patterns. *Mol Oncol* 2018; **12**(7): 1047-60.
19. Chen WB, Zhuang J, Wang PP, et al. DNA methylation-based classification and identification of renal cell carcinoma prognosis-subgroups. *Cancer Cell International* 2019; **19**.
20. Dong RZ, Yang X, Zhang XY, et al. Predicting overall survival of patients with hepatocellular carcinoma using a three-category method based on DNA methylation and machine learning. *J Cell Mol Med* 2019; **23**(5): 3369-74.

21. Li X, Cai Y. Better prognostic determination and feature characterization of cutaneous melanoma through integrative genomic analysis. *Aging (Albany NY)* 2019; **11**(14): 5081-107.

22. Wang Y, Deng H, Xin S, Zhang K, Shi R, Bao X. Prognostic and predictive value of three DNA methylation signatures in lung adenocarcinoma. *Frontiers in Genetics* 2019; **10**(APR).

23. Yang C, Zhang Y, Xu X, Li W. Molecular subtypes based on DNA methylation predict prognosis in colon adenocarcinoma patients. *Aging (Albany NY)* 2019; **11**(24): 11880-92.

24. Bian J, Long JY, Yang X, et al. Signature based on molecular subtypes of deoxyribonucleic acid methylation predicts overall survival in gastric cancer. *World J Gastroenterol* 2020; **26**(41): 6414-30.

25. Cai Q, He B, Xie H, et al. Identification of a novel prognostic DNA methylation signature for lung adenocarcinoma based on consensus clustering method. *Cancer Med* 2020; **9**(20): 7488-502.

26. Chen H, Ma X, Yang M, Wang M, Li L, Huang T. A methylomics-associated nomogram predicts recurrence-free survival of thyroid papillary carcinoma. *Cancer Med* 2020; **9**(19): 7183-93.

27. Deng Y, Wan H, Tian J, et al. CpG-methylation-based risk score predicts progression in colorectal cancer. *Epigenomics* 2020; **12**(7): 605-15.

28. Dong M, Yang Z, Li X, Zhang Z, Yin A. Screening of methylation gene sites as prognostic signature in lung adenocarcinoma. *Yonsei Medical Journal* 2020; **61**(12): 1013-23.

29. He H, Chen D, Cui SM, et al. HDNA methylation data-based molecular subtype classification related to the prognosis of patients with hepatocellular carcinoma. *Bmc Medical Genomics* 2020; **13**(1).

30. Lee TY, Huang KY, Chuang CH, Lee CY, Chang TH. Incorporating deep learning and multi-omics autoencoding for analysis of lung adenocarcinoma prognostication. *Computational Biology and Chemistry* 2020; **87**.

31. Li C, Ke J, Liu J, Su J. DNA methylation data-based molecular subtype classification related to the prognosis of patients with cervical cancer. *J Cell Biochem* 2020; **121**(3): 2713-24.

32. Li TD, Chen X, Gu ML, Deng AM, Qian C. Identification of the subtypes of gastric cancer based on DNA methylation and the prediction of prognosis. *Clinical Epigenetics* 2020; **12**(1).

33. Lian QX, Wang B, Fan LJ, Sun JQ, Wang GL, Zhang JD. DNA methylation data-based molecular subtype classification and prediction in patients with gastric cancer. *Cancer Cell International* 2020; **20**(1).

34. Luo H, Zhao Q, Wei W, et al. Circulating tumor DNA methylation profiles enable early diagnosis, prognosis prediction, and screening for colorectal cancer. *Sci Transl Med* 2020; **12**(524).

35. Luo R, Song J, Xiao X, et al. Identifying CpG methylation signature as a promising biomarker for recurrence and immunotherapy in non-small-cell lung carcinoma. *Aging (Albany NY)* 2020; **12**(14): 14649-76.

36. Lv J, Wang J, Shang X, Liu F, Guo S. Survival prediction in patients with colon adenocarcinoma via multiomics data integration using a deep learning algorithm. *Bioscience Reports* 2020; **40**(12).

37. Ma H, Zhao C, Zhao Z, et al. Specific glioblastoma multiforme prognostic-subtype distinctions based on DNA methylation patterns. *Cancer Gene Ther* 2020; **27**(9): 702-14.

38. Ma X, Chen H, Wang G, Li L, Tao K. DNA methylation profiling to predict overall survival risk in gastric cancer: development and validation of a nomogram to optimize clinical management. *J Cancer* 2020; **11**(15): 4352-65.

39. Macías-García L, Martínez-Ballesteros M, Luna-Romera JM, García-Heredia JM, García-Gutiérrez J, Riquelme-Santos JC. Autoencoded DNA methylation data to predict breast cancer recurrence: Machine learning models and gene-weight significance. *Artif Intell Med* 2020; **110**: 101976.

40. Shi S, Xu M, Xi Y. Molecular subtypes based on DNA promoter methylation predict prognosis in lung adenocarcinoma patients. *Aging (Albany NY)* 2020; **12**(23): 23917-30.

41. Tian ZJ, Meng LF, Long XB, et al. DNA methylation-based classification and identification of bladder cancer prognosis-associated subgroups. *Cancer Cell International* 2020; **20**(1).

42. Wang X, Wang D, Liu J, Feng M, Wu X. A novel CpG-methylation-based nomogram predicts survival in colorectal cancer. *Epigenetics* 2020; **15**(11): 1213-27.

43. Wang Y, Wang Y, Wang Y, Zhang Y. Identification of prognostic signature of non-small cell lung cancer based on TCGA methylation data. *Sci Rep* 2020; **10**(1): 8575.

44. Xiang R, Fu T. Gastrointestinal adenocarcinoma analysis identifies promoter methylation-based cancer subtypes and signatures. *Sci Rep* 2020; **10**(1): 21234.

45. Yu J, Wu X, Lv M, et al. A model for predicting prognosis in patients with esophageal squamous cell carcinoma based on joint representation learning. *Oncology Letters* 2020; **20**(6).

46. Zhang C, Wang F, Guo F, et al. A 13-gene risk score system and a nomogram survival model for predicting the prognosis of clear cell renal cell carcinoma. *Urologic Oncology: Seminars and Original Investigations* 2020; **38**(3): 74.e1-.e11.

47. Zhang E, Hou X, Hou B, Zhang M, Song Y. A risk prediction model of DNA methylation improves prognosis evaluation and indicates gene targets in prostate cancer. *Epigenomics* 2020; **12**(4): 333-52.

48. Zheng J, Zhang T, Guo W, et al. Integrative Analysis of Multi-Omics Identified the Prognostic Biomarkers in Acute Myelogenous Leukemia. *Frontiers in Oncology* 2020; **10**.

49. Bedon L, Dal Bo M, Mossenta M, Busato D, Toffoli G, Polano M. A Novel Epigenetic Machine Learning Model to Define Risk of Progression for Hepatocellular Carcinoma Patients. *Int J Mol Sci* 2021; **22**(3).

50. Chen H, Qin Q, Xu Z, et al. DNA methylation data-based prognosis-subtype distinctions in patients with esophageal carcinoma by bioinformatic studies. *Journal of Cellular Physiology* 2021; **236**(3): 2126-38.

51. Hao XY, Li AQ, Shi H, et al. A novel DNA methylation-based model that effectively predicts prognosis in hepatocellular carcinoma. *Bioscience Reports* 2021; **41**(3).

52. Huang G, Zhang J, Gong L, et al. Specific Lung Squamous Cell Carcinoma Prognosis-Subtype Distinctions Based on DNA Methylation Patterns. *Med Sci Monit* 2021; **27**: e929524.

53. Huang GJ, Wang C, Fu X. Bidirectional deep neural networks to integrate RNA and DNA data for predicting outcome for patients with hepatocellular carcinoma. *Future Oncology* 2021.

54. Huang H, Fu JM, Zhang L, et al. Integrative Analysis of Identifying Methylation-Driven Genes Signature Predicts Prognosis in Colorectal Carcinoma. *FRONTIERS IN ONCOLOGY* 2021; **11**.

55. Huang W, Weng W, Wu B, et al. Development and validation of the trans-omics model for pancreatic adenocarcinoma. *Epigenomics* 2021; **13**(1): 15-30.

56. Lee N, Xia X, Meng H, et al. Identification of a novel CpG methylation signature to predict prognosis in lung squamous cell carcinoma. *Cancer Biomarkers* 2021; **30**(1): 63-73.

57. Li K, Wu Z, Yao J, Fan J, Wei Q. DNA methylation patterns-based subtype distinction and identification of soft tissue sarcoma prognosis. *Medicine (Baltimore)* 2021; **100**(5): e23787.

58. Li XS, Nie KC, Zheng ZH, et al. Molecular subtypes based on DNA methylation predict prognosis in lung squamous cell carcinoma. *BMC Cancer* 2021; **21**(1): 96.

59. Peng YJ, Zhao J, Yin F, et al. A methylation-driven gene panel predicts survival in patients with colon cancer. *Febs Open Bio* 2021; **11**(9): 2490-506.

60. Shang SP, Li X, Gao Y, et al. MeImmS: Predict Clinical Benefit of Anti-PD-1/PD-L1 Treatments Based on DNA Methylation in Non-small Cell Lung Cancer. *Frontiers in Genetics* 2021; **12**.

61. Sun GP, Duan H, Xing YH, Zhang DW. Prognostic Score Model Based on Ten Differentially Methylated Genes for Predicting Clinical Outcomes in Patients with Adenocarcinoma of the Colon. *CANCER MANAGEMENT AND RESEARCH* 2021; **13**: 5113-25.

62. Tan S, Gui WW, Wang SM, Sun CQ, Xu X, Liu LX. A methylation-based prognostic model predicts survival in patients with colorectal cancer. *JOURNAL OF GASTROINTESTINAL ONCOLOGY* 2021; **12**(4): 1590-600.

63. Wang L, Xue FX. Prognosis-associated methylation subtypes in endometrial carcinoma patients and the role of magnetic nanoparticles in gene extraction. *Materials Express* 2021; **11**(8): 1288-98.

64. Wang TH, Lee CY, Lee TY, Huang HD, Hsu JBK, Chang TH. Biomarker Identification through Multiomics Data Analysis of Prostate Cancer Prognostication Using a Deep Learning Model and Similarity Network Fusion. *Cancers* 2021; **13**(11).

65. Wu ZH, Tang Y, Zhou Y. DNA Methylation Based Molecular Subtypes Predict Prognosis in Breast Cancer Patients. *Cancer Control* 2021; **28**.

66. Xu QH, Hu YB, Chen SH, et al. Immunological Significance of Prognostic DNA Methylation Sites in Hepatocellular Carcinoma. *Frontiers in Molecular Biosciences* 2021; **8**.

67. Yin LL, Zhang NN, Yang Q. DNA methylation subtypes for ovarian cancer prognosis. *Febs Open Bio* 2021; **11**(3): 851-65.

68. Yin XL, Kong LM, Liu P. Identification of prognosis-related molecular subgroups based on DNA methylation in pancreatic cancer. *Clinical Epigenetics* 2021; **13**(1).

69. Yu X, Yang Q, Wang D, Li Z, Chen N, Kong DX. Predicting lung adenocarcinoma disease progression using methylation-correlated blocks and ensemble machine learning classifiers. *PeerJ* 2021; **9**.
